# THE STATE OF CURRENT MANAGEMENT OF THE HEIGHTENED RISK FOR ATHEROSCLEROTIC CARDIOVASCULAR EVENTS IN AN ESTABLISHED COHORT OF PATIENTS WITH LUPUS ERYTHEMATOSUS

**DOI:** 10.1101/2022.12.12.22283349

**Authors:** Megan Zhao, Rui Feng, Victoria P. Werth, Kevin Jon Williams

## Abstract

**Background:** Patients with lupus erythematosus (LE) are at a heightened risk for clinical events, chiefly heart attacks and strokes, caused by atherosclerotic cardiovascular disease (ASCVD). We recently proposed new guidelines to categorize levels of risk for future ASCVD events specifically in LE patients, with recommendations for management. Here, we assessed the state of current management of ASCVD event risk in light of these new recommendations.

**Methods:** We studied our entire UPenn Longitudinal Lupus Cohort of patients with cutaneous LE, without or with concurrent systemic LE, for whom we had full access to medical records (n=370, LE-ASCVD Study Cohort, years 2007-2021).

**Results:** Of our LE-ASCVD Study Cohort, 336/370 (90.8%) had a designated primary-care physician. By the new guidelines, the most recent plasma LDL cholesterol levels were above goal for 252/370 (68.1%) of the Cohort. Two hundred sixty-six (71.9%) had hypertension, which was under- or un-treated in 198/266 (74.4%). Of current smokers, 51/63 (81.0%) had no documented smoking cessation counseling or referrals. Diabetes was generally well-managed, and hypertriglyceridemia was uncommon. Of the Cohort, 254 patients qualified for two widely used online calculators in primary prevention that estimate the risk of an ASCVD event in the next 10 years: the ACC-ASCVD Risk Estimator Plus and QRisk3. We also stratified these 254 patients into the categories of ASCVD event risk we recently defined specifically for LE. Surprisingly, these three methods for estimating ASCVD event risk showed clinically meaningful agreement for only 100/254 (39.4%), i.e., discordance for over 60% that could affect clinical management. The documented rate of ASCVD events in the first 10 years after enrollment was 22.3% (95% CI 16.9%, 27.4%), indicating a high-risk population despite a preponderance of women and a median age at enrollment of only 47 years.

**Conclusion:** Cutaneous LE patients are under-treated compared with the new guidelines and, accordingly, they experience a substantial burden of ASCVD events. Moreover, it is unclear how to accurately assess future ASCVD event risk in these patients – except that it is high – and this uncertainty may complicate clinical management. Efforts are underway to improve ASCVD event risk estimation and guideline implementation in lupus patients.

## Introduction

Patients with lupus erythematosus (LE) suffer from a heightened risk of clinical events, chiefly heart attacks and strokes, caused by atherosclerotic cardiovascular disease (ASCVD). Several clinical features of LE contribute to the problem.[1] First, patients with LE exhibit an increased prevalence of conventional risk factors for ASCVD events, particularly dyslipoproteinemia and hypertension, and glucocorticoid use often exacerbates additional conditions related to cardiovascular risk, such as obesity, the metabolic syndrome, so-called “prediabetes” (defined as dysglycemia to an extent that indicates a high risk of developing diabetes), and type 2 diabetes mellitus.[1, 2] Moreover, patients with cutaneous LE (CLE) smoke at higher rates than in the general population.[3–5] Second, numerous studies using arterial ultrasonography, coronary CT angiography, and other methods have shown that LE patients carry an increased burden of atherosclerotic plaques compared with the general population.[1, 6–9] Third, the increased rate of ASCVD events amongst LE patients persists even after adjustment for age and sex and, more strikingly, even after adjustment for other conventional risk factors for ASCVD events.[1, 10, 11] We have suggested that autoimmune processes specific to LE may exacerbate specific, known pathogenic steps in atherosclerosis.[1]

Despite their high plaque burden and heightened risk for ASCVD events, however, LE patients remain undertreated for the causative agents of ASCVD, meaning low-density lipoprotein (LDL) and other cholesterol-rich apolipoprotein-B (apoB)-containing lipoproteins,[12, 13] and key exacerbators of ASCVD, such as hypertension, smoking, and hyperglycemia.[1, 14–17] As part of our efforts to address this problem, we recently proposed a system for assessing CLE and systemic LE (SLE) patients using four defined categories of ASCVD event risk, with corresponding guidance for clinical management of LE patients in each ASCVD event risk category using lifestyle modifications and modern medications.[1]

In the current work, we performed a single-center study of all 370 patients with CLE, without or with concurrent SLE, within our established longitudinal cohort at the University of Pennsylvania Health System (UPHS) for whom we have full access to their medical records.[18–23] Our goal was to assess the state of current management of ASCVD event risk through conventional therapeutic targets, chiefly hyperlipidemia, hypertension, smoking, prediabetes, and diabetes mellitus, in the light of the new recommendations.[1] We also studied two earlier and still widely used online tools for estimating the risk of an ASCVD event in the next 10 years (“10-year ASCVD event risk”) for individuals without clinically evident ASCVD (primary prevention) from the general population, i.e., not specifically designed for CLE patients: the American College of Cardiology (ACC) ASCVD Risk Estimator Plus[24, 25] and the QRisk3 calculator.[26–28]

## Methods

### The LE-ASCVD Study Cohort

We previously reported the UPenn Longitudinal Lupus Cohort, which is our established cohort of patients receiving care at UPHS whom we have been recruiting from our specialty Dermatology clinic since 2007 based on a diagnosis of CLE, without or with concurrent SLE.[18–23] The UPHS Institutional Review Board approved patient recruitment and then collection and analyses of data, and we obtained written informed consent from each participant. This study followed the principles outlined in the Declaration of Helsinki.

For the current study, we required full access to longitudinal and recent medical records. Thus, we included only active UPHS patients and patients whose records from outside hospitals were either linked or faxed into the UPHS electronic medical record (“LE-ASCVD Study Cohort”). Our database consisted of clinical information for these patients, including ASCVD events and conventional parameters relevant to management of future ASCVD event risk, that we entered into the University’s secure RedCap system (Research Electronic Data Capture, Vanderbilt University, Nashville, TN, USA) to ensure compliance with confidentiality requirements of the Health Insurance Portability and Accountability Act of 1996 (HIPAA).

### Collection and interpretation of clinical data

Our approaches for collecting and interpreting these data are described in detail in the Supplementary Methods, in the sections, *Demographic and clinical data collected for the RedCap database* and *Three systems to estimate the risk of a future ASCVD event for individual patients within the LE-ASCVD Study Cohort*.

Among the conventional parameters relevant to future ASCVD event risk, the lipid/lipoprotein panels in the patients’ electronic charts uniformly included plasma concentrations of total cholesterol (TC), LDL cholesterol (LDLc), triglycerides (TG), and high-density lipoprotein cholesterol (HDLc), but not apolipoprotein-B (apoB) nor a one-time assay for lipoprotein(a) [Lp(a)].[1, 29–31] Non-HDLc, a convenient parameter that captures the cholesterol carried by LDL and all other atherogenic apoB-containing lipoproteins, was calculated as TC minus HDLc.

All previously documented or newly incident major adverse atherosclerotic cardiovascular events (MAACE) were noted along with the event date and were defined as atherosclerotic myocardial infarction (MI), atherosclerotic ischemic heart disease other than MI such as angina and coronary revascularization, atherosclerotic (non-embolic) ischemic stroke or transient cerebrovascular ischemic attack, newly diagnosed symptomatic peripheral arterial disease (PVD), or hospitalization for chest pain, shortness of breath, or palpitations with suspicion of ischemic origin. Major adverse cardiovascular events (MACE) that were clearly not atherosclerotic were excluded, such as coronary vasospasm without documentation of nearby plaque, venous thrombosis or pulmonary embolism, embolic stroke unrelated to atherosclerosis, and heart failure from non-ischemic causes (chronic hypertension, diabetes mellitus, non-ischemic valvular disorders, post-myocarditis, and congenital or genetic defects).

### Statistical analyses

Categorical parameters are given as n (%). Continuous variables are given as mean±SD, if normally distributed, or as median (interquartile range, IQR), if non-normally distributed.

To assess the correlation between the numerical estimates of 10-year ASCVD event risk from the QRisk3 versus the ACC Risk Estimator Plus for all patients in the LE-ASCVD Study Cohort for whom we could use both, we calculated a Pearson’s correlation coefficient (r) and fit a linear regression. Documented MAACE were tabulated for the period before enrollment into the Cohort and then for the entire period of study after enrollment. Event-free survival rates since enrollment in the LE-ASCVD Study Cohort were computed according to Kaplan & Meier, along with 95% confidence intervals.[32, 33]

## Results

The UPenn Longitudinal Lupus Cohort currently consists of 529 CLE patients.[21–23] Upon review, we found that 370 of them have electronic medical records that are fully accessible within UPHS and therefore could be included in our LE-ASCVD Study Cohort.

Table 1 shows demographics and other characteristics of the LE-ASCVD Study Cohort. Consistent with our previous reports,[1, 21, 23] the LE-ASCVD Study Cohort was predominantly female (82.4%), with nearly nine out of 10 identifying as White (51.4%) or Black (38.1%), consistent with the UPHS catchment area. Average age was 47.1±14.9 years at enrollment and 54.8±15.9 years at the end of the data collection period (Table 1). Average length of time since enrollment into the UPenn Longitudinal Lupus Cohort was 7.7±4.5 years.

**Table 1.** Demographics and other characteristics of the LE-ASCVD Study Cohort. Here and elsewhere, categorical parameters are given as n (%). Continuous variables are given as mean±SD, if normally distributed, or as median (interquartile range, IQR), if non-normally distributed.

| Category | Parameter | Value | n in LE-ASCVD Study Cohort or subgroup |
| --- | --- | --- | --- |
| **Demographics** |  |  |  |
|  | Female | 305 (82.4%) | 370 (entire LE-ASCVD Study Cohort) |
|  | White | 190 (51.4%) | 370 |
|  | Black | 141 (38.1%) | 370 |
|  | Asian | 15 (4.1%) | 370 |
|  | Hispanic | 17 (4.6%) | 370 |
| **Clinical course and care in the Cohort** |  |  |  |
|  | Age at enrollment into the Cohort | 47.1±14.9 years | 370 |
|  | Age as of 2021 | 54.8±15.9 years | 370 |
|  | Length of time since enrollment into the Cohort | 7.7±4.5 years | 370 |
|  | Extreme Risk for future ASCVD events* | 82 (22.2%) | 370 |
|  | CLE patients with concurrent SLE | 190 (51.4%) | 370 |
|  | History of lupus nephritis | 42 (11.4%) | 370 |
|  | History of CKD stage 3 or 4‡ | 37 (10.0%) | 370 |
|  | Hypertension§ | 266 (71.9%) | 370 |
|  | Former Smokers | 109 (29.5%) | 370 |
|  | Current Smokers | 63 (17.0%) | 370 |
|  | Most recent BMI | 28.4±7.1 kg/m<sup>2</sup> | 346 (subgroup with at least one set of simultaneous height and weight measurements) |
|  | Obese (BMI ≥30.0 kg/m<sup>2</sup>) | 116 (33.5%) | 346 |
|  | Prediabetes** | 67 (18.1%) | 370 |
|  | Diabetes mellitus** | 39 (10.5%) | 370 |
| **Provider Data** |  |  |  |
|  | Patients with a listed PCP in the chart | 336 (90.8%) | 370 |
|  | Patients with documented PCP visit in the chart | 279 (75.4%) | 370 |

BMI, body mass index; PCP, primary care provider.
\*Extreme ASCVD event risk was defined by Keyes et al. 2021 as “Patients with LE ... and clinically evident ASCVD”, i.e., secondary prevention in the context of LE.<sup>[1]</sup>
‡An additional five (1.4%) patients in the LE-ASCVD Study Cohort were in CKD stage 5, meaning an eGFR below 15 ml/min/1.73m<sup>2</sup>.
§Hypertension was defined following the recently updated joint guidelines from the ACC and the American Heart Association (AHA),<sup>[34]</sup> here based on averaged blood pressure readings from the most recent year of follow-up, plus the list of active prescriptions in the electronic charts. Three hundred forty-one patients (92.3% of the LE-ASCVD Cohort) had two or more readings in their electronic charts in this period; 28 patients (7.6%) had only one reading; and 1 patient had no available blood pressure value in the most recent year.
\*\*Following criteria from the American Diabetes Association (ADA), HbA<sub>1c</sub> values of 5.7-6.4% indicate prediabetes, and HbA<sub>1c</sub> ≥6.5% is diagnostic for diabetes mellitus.<sup>[34]</sup> Accordingly, prediabetes was defined here as chart-diagnosed or the most recent HbA<sub>1c</sub> was 5.7-6.4%, without a diagnosis of overt diabetes on the chart. Diabetes was defined as chart-diagnosed or the most recent HbA<sub>1c</sub> was ≥6.5%. The most recent HbA<sub>1c</sub> value for some patients with a diagnosis of diabetes mellitus was below 6.5%; we still classified them under diabetes unless a clinician had marked the diagnosis on the chart as resolved or in remission.<sup>[34-38]</sup>

As of their most recent medical encounter, 22.2% of the patients in our LE-ASCVD Study Cohort had clinically evident ASCVD, meaning a diagnosis of MAACE as defined in the Methods, which classifies these individuals as candidates for secondary prevention (n=82/370; Extreme Risk in Table 1 and in Figure 1A; no patient qualified for the highest-risk category from Keyes et al. of Recent Recurrent ASCVD Events). Just over half of the LE-ASCVD Study Cohort had concurrent SLE (51.4%), with 11.4% having a history of lupus nephritis, and 10.0% having a history of stage 3 or 4 CKD. Median times since diagnosis of CLE or concurrent SLE were over a decade (Supplementary Table S1).

**Figure 1.**
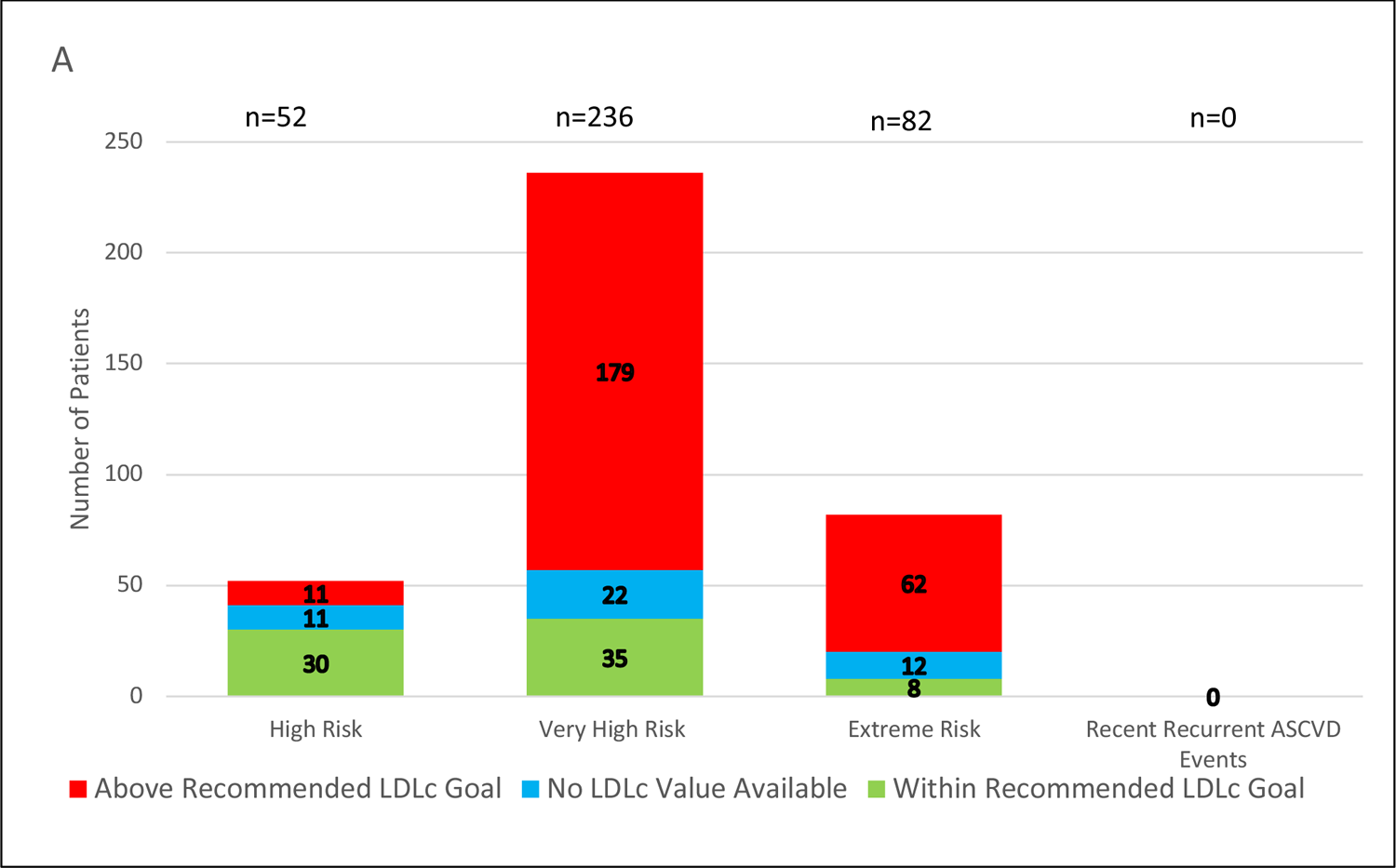
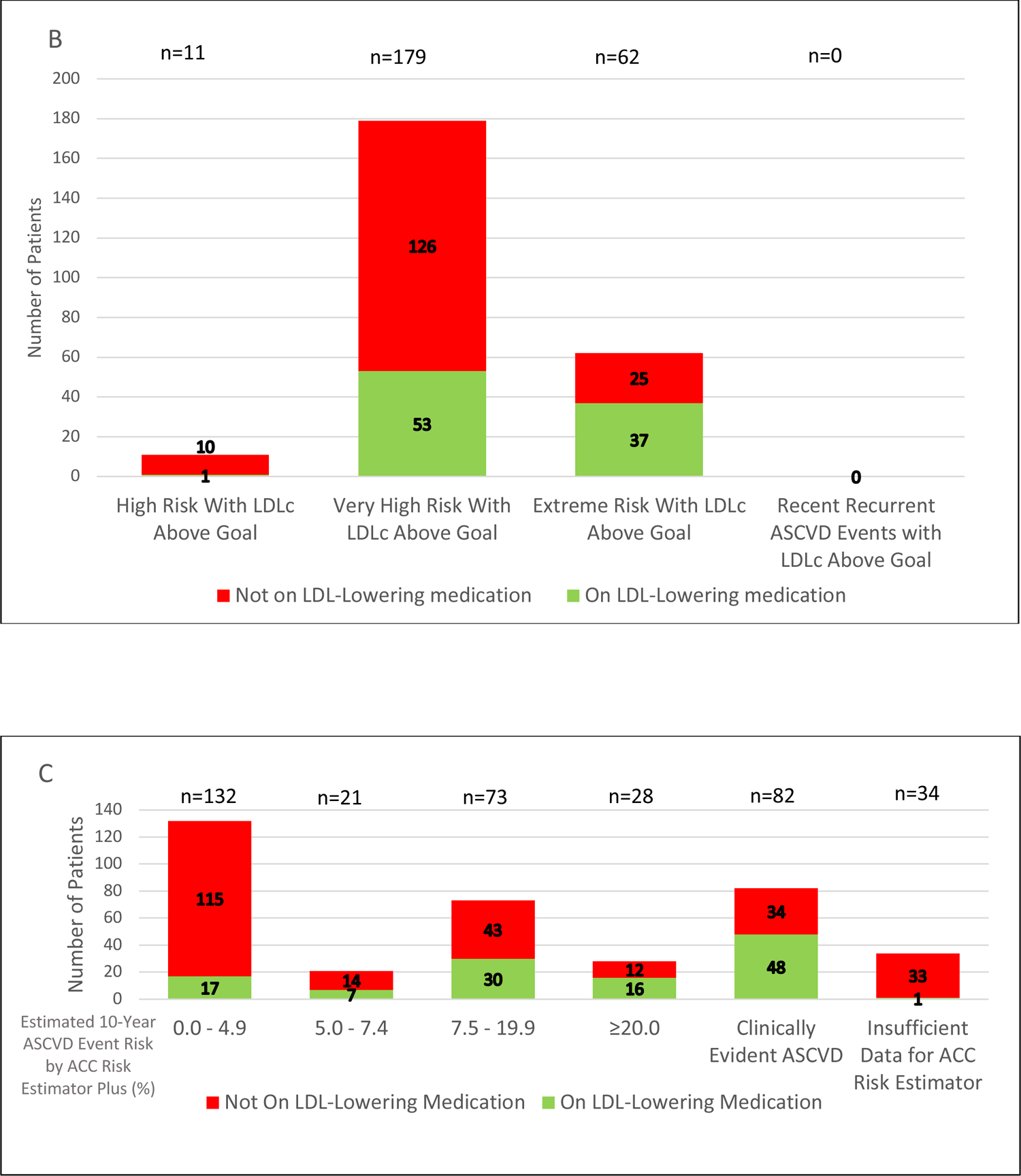
Assessment and management of plasma low-density lipoprotein cholesterol (LDLc) concentrations in the LE-ASCVD Study Cohort. <u>Panel A</u>: Classification of Study Cohort patients into the four categories of ASCVD event risk defined by the newly proposed guidelines of Keyes et al. for lupus patients.[1] Also indicated are numbers of patients with LDLc levels above (red) or within (green) the newly proposed goals for each risk category. Patients without an available LDLc value are indicated in blue. <u>Panel B</u>: Absence (red) or presence (green) of an active prescription for an LDL-lowering medication in the electronic charts of patients classified into each of the four newly proposed categories of ASCVD event risk for lupus patients and whose most recent plasma LDLc levels were above the newly proposed goals; these are the patients indicated in red in panel 1A. <u>Panel C</u>: Estimated risk of an ASCVD event in the next 10 years (“10-year ASCVD event risk”) by the online ACC Risk Estimator Plus and absence (red) or presence (green) of an active prescription for an LDL-lowering medication in the electronic chart. Patients are grouped according to the cut-offs for estimated 10-year ASCVD event risk from the ACC/AHA guidelines for primary prevention of cardiovascular disease in the general population (0.0-4.9%, 5.0-7.4%, &c.).[25] As noted in the Supplementary Methods, patients below age 40 years without clinically evident ASCVD for whom we had sufficient data for the ACC ASCVD Risk Estimator Plus (n=47/370) were scaled up to 40, and patients above age 79 years without clinically evident ASCVD (n=13/370) were scaled down to 79. Data are presented with these additional patients included (here) or presented separately in their own columns (Supplementary Figure S1C). Also shown here are data for patients with clinically evident ASCVD (meaning secondary prevention, for which the ACC ASCVD Risk Estimator Plus does not apply), as well as data for patients with no clinically evident ASCVD but insufficient information in the electronic chart for the ACC Risk Estimator (rightmost column). At the top of each column in panels A-C is indicated the total number of patients represented by that column. Numbers of patients represented by each tinted portion of each column are also given.

Almost three-quarters of the patients in the LE-ASCVD Study Cohort (71.9%, Table 1) had hypertension, defined as a systolic blood pressure ≥130, a diastolic pressure ≥80 mm Hg, and/or on anti-hypertensive medication, following the recently updated joint guidelines from the ACC and the American Heart Association (AHA).[17, 34] Of the Study Cohort, 29.5% were former smokers, and 17.0% are current smokers (Table 1). Of the subgroup with at least one set of simultaneous height and weight measurements, approximately one-third were underweight or normal weight (n=125/346, 36.2%), one-third overweight (30.3%), and one-third obese (33.5%; Table 1 and Supplementary Table S1). Prediabetes was present in 18.1%, and diabetes in 10.5%, of the LE-ASCVD Study Cohort (Table 1). A diagnosis of depression was common (37.6%; Supplementary Table S1), as previously reported for CLE.[35]

Regarding management, nearly all the patients in the LE-ASCVD Study Cohort had a primary-care physician (PCP) listed in the electronic medical record (90.8%; Table 1). Time elapsed since the last visit with a primary-care physician was 10 months (IQR 4.0-31; Supplementary Table S1). Number of physician providers, excluding trainees, was 5.8±2.8 per patient (mean±SD; Supplementary Table S1). Most patients in the Study Cohort had seen a rheumatologist (203/307, 54.9%), and under half had seen a cardiologist (40.8%), nephrologist (15.4%), or endocrinologist (8.0%; Supplementary Table S1).

### Assessments and management of lipid/lipoprotein levels as targets to lower ASCVD event risk

The vast majority (n=325/370, 87.8%) of patients in the LE-ASCVD Study Cohort had plasma lipid/lipoprotein levels checked at least once, with a median elapsed time of 35 months (IQR 14-83) since the most recent panel, i.e., almost three years (Supplementary Table S1). We found that PCPs checked just over 60% of the most recent lipid/lipoprotein values, and non-PCPs checked the rest. Of the non-PCPs, rheumatologists checked lipid/lipoprotein values in more patients than did other subspecialists (Supplementary Table S1).

Based on data from the electronic charts, including absence (primary prevention) or presence (secondary prevention) of clinically evident ASCVD, as well as major conventional risk factors for ASCVD events, we classified patients from the LE-ASCVD Cohort into the four defined categories of ASCVD event risk from our recently proposed guidelines for LE patients – namely, High Risk, Very High Risk, Extreme Risk, and Recent Recurrent ASCVD Events.[1] Next, we compared current clinical care with the new therapeutic goals for each of these categories of ASCVD event risk for LE patients.[1]

We found that almost one-quarter of LE patients classified at High Risk for an ASCVD event had plasma LDLc concentrations that were above the newly recommended ranges (n=11/52, 21.2%; red in the leftmost column of Figure 1A). More strikingly, over three-quarters of LE patients classified at either Very High Risk (179/236, 75.8%) or Extreme Risk (62/82, 75.6%) for an ASCVD event also had plasma LDLc concentrations above the newly recommended ranges (red in the middle two columns of Figure 1A). For the 252 LE patients whose most recent LDLc value was above-goal, the time elapsed since that above-goal LDLc was 47 months (IQR 24-97), i.e., almost four years (Supplementary Table S1). Data for non-HDLc levels were similar: 13.5% (7/52), 66.5% (157/236), and 72.0% (59/82) of our LE patients in these three categories of ASCVD event risk had values above the newly recommended ranges (Supplementary Figure S1A).

Regarding lipid/lipoprotein management in primary prevention, 90.9% (n=10/11) of High Risk patients with out-of-range LDLc levels were not on any LDL-lowering medications recorded in their charts, and 70.4% (n=126/179) of Very High Risk patients with out-of-range LDLc levels were not on LDL-lowering medications (Figure 1B). Again, data for non-HDLc levels were similar: 85.7% (6/7), and 72.6% (114/157) of LE patients in these two categories of ASCVD event risk with out-of-range non-HDLc levels were not on any LDL-lowering medications (Supplementary Figure S1B). Thus, over two-thirds of LE patients classified at High or Very High ASCVD event risk with out-of-range LDLc or non-HDLc values according to the newly published guidelines were on no LDL-lowering medications.

Of the 82 LE patients classified at Extreme ASCVD event risk (secondary prevention), only about one in ten achieved plasma concentrations of LDLc (8/82, 9.8%) or non-HDLc (11/82, 13.4%) within the new goals. Nearly half were under-treated by the new guidelines, meaning on LDL-lowering medications but still with out-of-range values for LDLc (37/82, 45.1%) or non-HDLc (34/82, 41.5%). About 30% were untreated, meaning out-of-range values for LDLc (25/82, 30.5%) or non-HDLc (also 25/82, 30.5%) yet on no LDL-lowering medications. The remainder of the LE patients classified at Extreme ASCVD event risk had no LDLc (12/82, 14.6%) or HDLc (also 12/82, 14.6%) values available from their charts despite documentation of clinically evident ASCVD, the key requirement for this risk category[1] (Figure 1B, Supplementary Figure S1B).

We used the ACC ASCVD Risk Estimator Plus[24, 25] to estimate the risk of an ASCVD event in the next 10 years (“10-year ASCVD event risk”) for all patients in the LE-ASCVD Study Cohort meeting the criteria of no clinically evident ASCVD (primary prevention, n=288/370, 77.8% of the Study Cohort; Figure 1A, Table 1), with sufficient available clinical data to allow use of this online risk estimator (n=254), and ages 40-79 years, a stated requirement for this online calculator to estimate 10-year ASCVD event risks (n=194). By performing age-scaling as described in the Supplementary Methods, we included an additional 47 patients below the age of 40 years (12.7% of the LE-ASCVD Study Cohort) and 13 patients above 79 years (3.5% of the Study Cohort) without clinically evident ASCVD and with sufficient data for this online ASCVD event risk estimator, to analyze the entire subset of 254 patients just mentioned, i.e., nearly 90% of the LE-ASCVD Study Cohort who had no clinically evident ASCVD (254/288, 88.2%).

Figure 1C displays data for these 254 patients stratified by estimates of their 10-year ASCVD event risk from the ACC ASCVD Risk Estimator Plus, following the cutoffs of Arnett et al. in the ACC/AHA guidelines for primary prevention of cardiovascular disease in the general population – namely, 0.0-4.9% 10-year ASCVD event risk (low), 5.0-7.4% (borderline), 7.5-19.9% (intermediate), and ≥20% (high).[25] Of the patients with an estimated 10-year ASCVD event risk of 7.5% to 19.9% from the ACC ASCVD Risk Estimator Plus, over half were not on any LDL-lowering medication (n=43 out of 73, 58.9%; Figure 1C). Of patients with an estimated 10-year ASCVD event risk ≥20%, 42.9% (n=12/28) were not on an LDL-lowering medication recorded in the chart. Supplementary Figure S1C shows nearly identical results from the original subset of 194 LE patients with no clinically evident ASCVD, sufficient data for this online risk estimator, and aged 40-79 years with no age-scaling.

Of the LE patients who already have clinically evident ASCVD and are therefore candidates for secondary prevention of ASCVD events, 41.5% (n=34/82) were not on any LDL-lowering medication; this figure comprises Extreme Risk patients on no LDL-lowering medication who have either (i) no LDLc value available (7 out of 12 from Figure 1A), (ii) the most recent LDLc value within goal (2 out of 8 from Figure 1A), or (iii) the most recent LDLc value above goal (25 from Figure 1B).

Supplementary Figure S1D shows data for patients in the LE-ASCVD Study Cohort stratified by their most recent plasma LDLc concentrations, and then within each LDLc concentration range, we indicate treatment, or not, with an LDL-lowering medication. In each LDLc concentration range, except for the lowest (<70 mg/dl) and the highest (≥190 mg/dl), over 60% of our LE patients were on no LDL-lowering medications. There was no LDLc value available in the chart for 45 patients (12.2% of the entire LE-ASCVD Study Cohort), nearly all of whom were on no LDL-lowering medications (n=39/45, 86.7%). Supplementary Figure S1E shows data from stratifying LE patients by their most recent plasma non-HDLc concentrations. Similar to our findings with LDLc, in each concentration range of non-HDLc, except for the lowest (<80 mg/dl) and the highest (≥220 mg/dl), over 60% of LE patients were on no LDL-lowering medications (Supplementary Figure S1E).

### Assessments and management of hypertension

As noted above, almost three-quarters of the LE-ASCVD Study Cohort had hypertension (266/370, 71.9%; Table 1). Of these 266 hypertensive lupus patients, approximately one-fourth were well-managed, meaning normotensive on anti-hypertensive medications (68/266, 25.6%; green in the leftmost column of Figure 2A); almost one half were under-treated, meaning on anti-hypertensive medications but still hypertensive (122/266, 45.9%; green in the middle column of Figure 2A); and almost one third were untreated, meaning hypertensive yet on no anti-hypertensive medication (76/266, 28.6%; red in the middle column of Figure 2A). These data indicate that hypertension is a common but undermanaged problem in lupus patients, consistent with recently published literature using these blood pressure cut-offs.[17]

**Figure 2.**
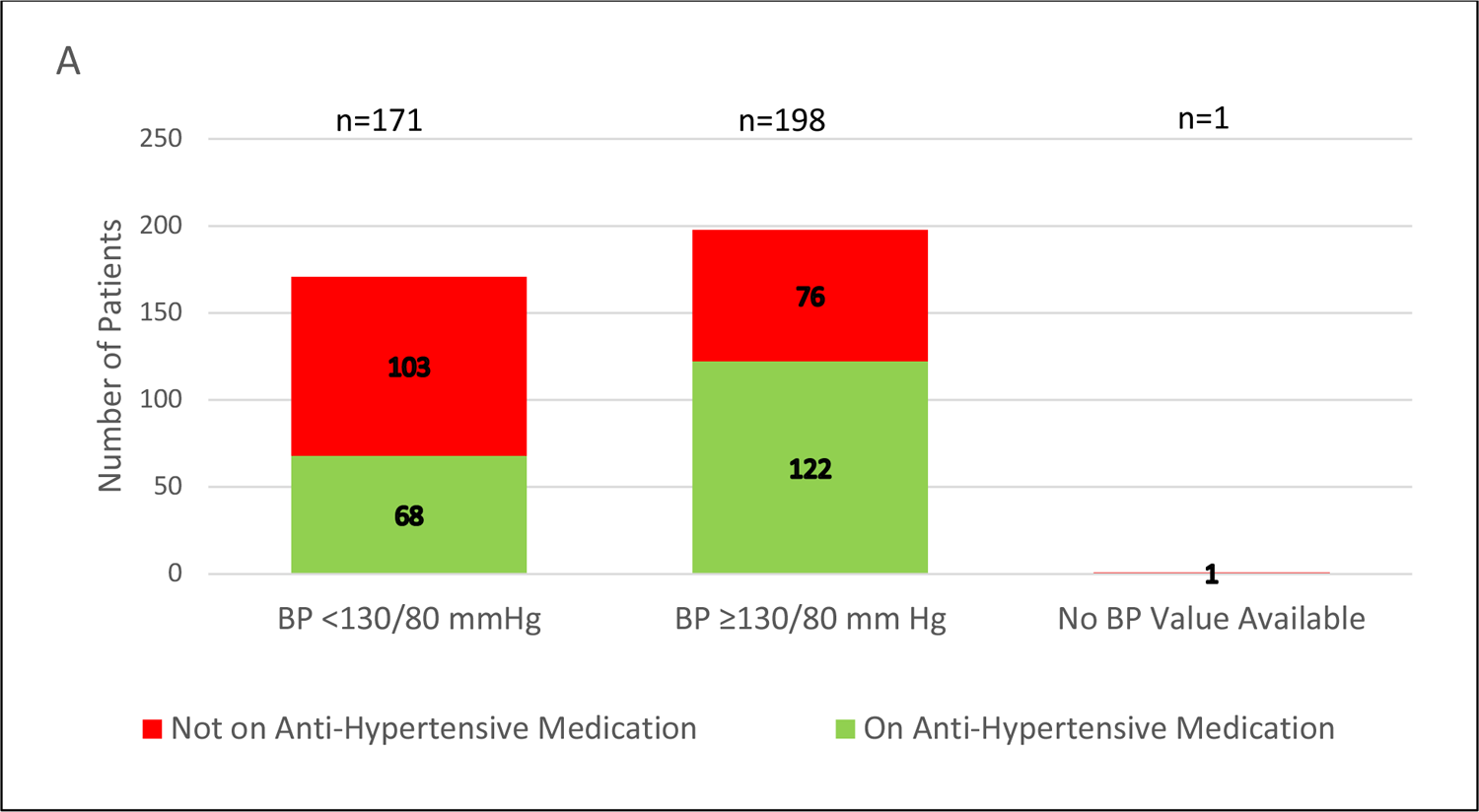
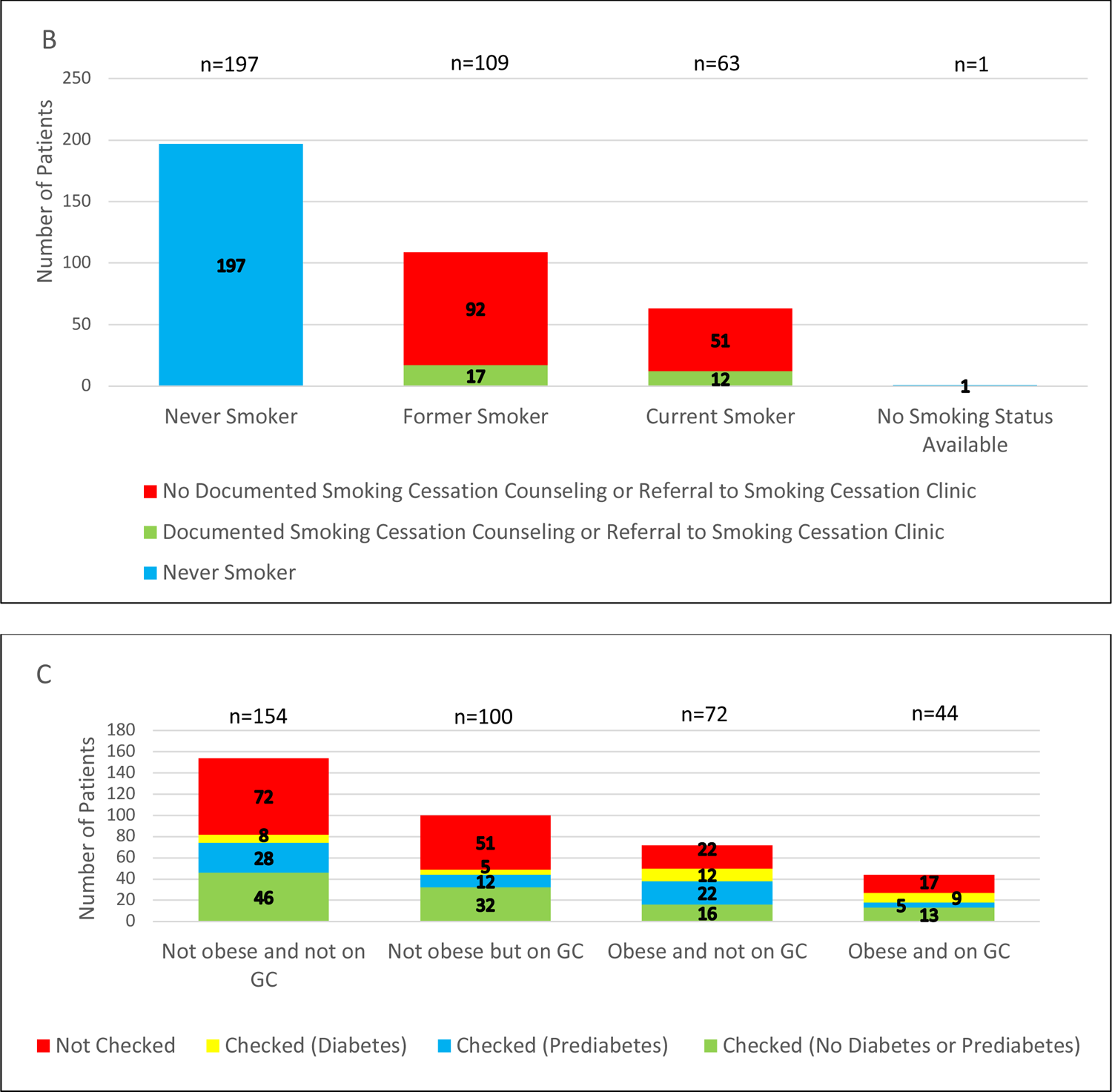
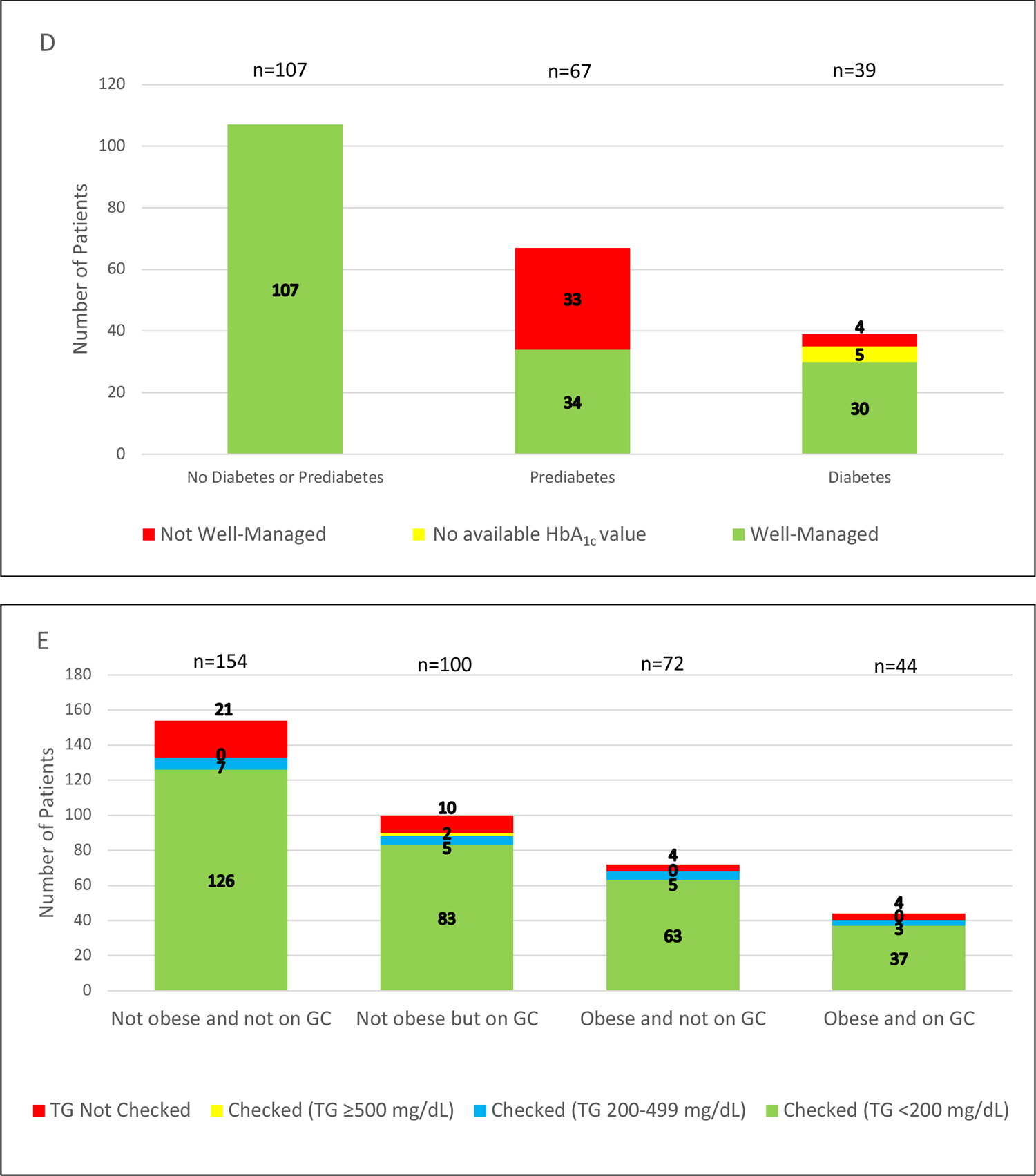
Assessment and management of several exacerbators and an additional causative agent of ASCVD event risk in the LE-ASCVD Study Cohort. <u>Panel A</u>: Prevalence and management of hypertension. Under-treated or untreated hypertension was defined by a systolic blood pressure (BP) ≥130 and/or a diastolic BP ≥80 mm Hg, indicated here as “BP≥130/80 mm Hg”. Red indicates patients with no record of an active prescription for an anti-hypertensive medication in the electronic chart; green indicates patients on at least one anti-hypertensive medication. <u>Panel</u> <u>B</u>: Prevalence and management of tobacco smoking. Red indicates patients with no chart documentation of either smoking cessation counseling or a referral to a smoking cessation clinic; green indicates patients with a record of one or both of these interventions. Data from never-smokers are also shown (blue). <u>Panel C</u>: Assessment, or not (red), of glycated hemoglobin (HbA_1c_) values for patients stratified by obesity and/or glucocorticoid (GC) use, two common factors in lupus patients that impair glycemic control. Diabetes was defined as in Table 1 as chart-diagnosed or the most recent HbA_1c_ was ≥6.5%; here, yellow indicates diabetes and at least one HbA_1c_ value on the chart. Five patients had a diagnosis of diabetes on their charts but no available HbA_1c_ value; these patients are included within the red portions of the columns here. Prediabetes (blue) was defined as chart-diagnosed or the most recent HbA_1c_ was 5.7-6.4%, without a diagnosis of overt diabetes on the chart. Green indicates patients with at least one HbA_1c_ value on the chart and no indication of either prediabetes or diabetes. <u>Panel D</u>: Management of prediabetes and diabetes in the LE-ASCVD Study Cohort. For prediabetes, well-managed (green) was defined here as documentation in the chart of lifestyle counseling and/or an active prescription for metformin. For diabetes, well-managed (green) was defined here as the most recent HbA_1c_ ≤7%.[36–40] Red indicates not well-managed by these definitions. Yellow in this panel indicates a diagnosis of diabetes but no available HbA_1c_ value on the chart. <u>Panel E</u>: Assessment, or not (red), of plasma triglyceride (TG) concentrations for patients stratified by obesity and/or GC use, two common factors in lupus patients that can elevate plasma TGs. Most recent plasma TG values are classified as very high (≥500 mg/dl, yellow), high (200-499 mg/dl, blue), or <200 mg/dl (green).

### Smoking status and management

Figure 2B indicates the prevalence and management of tobacco smoking in the LE-ASCVD Study Cohort. Of the former smokers in our cohort, the overwhelming majority quit smoking without any documentation in the chart of smoking cessation counseling or referral to a specialized smoking cessation clinic (n=92/109, 84.4%). Similarly, the overwhelming majority of current smokers have no documentation in their charts of smoking cessation counseling or clinic referral (n=51/63, 81.0%).

### Assessments and management of glycated hemoglobin (HbA_1c_) and plasma triglyceride levels

The majority (n=208/370, 56.2%) of patients in the LE-ASCVD Study Cohort had their HbA_1c_ levels checked at least once, with a median time elapsed time of 36 months (IQR 11-68) since the last assay, i.e., three years (Supplementary Table S1). As with lipid/lipoprotein values, PCPs checked just over 60% of the most recent HbA_1c_ levels, and non-PCPs checked the rest. Of the non-PCPs, rheumatologists checked HbA_1c_ levels in more patients than did other subspecialists (Supplementary Table S1).

Figure 2C shows assessment, or not, of HbA_1c_ values in the LE-ASCVD Study Cohort after stratification by obesity and/or current or previous glucocorticoid use, two common factors in lupus patients that impair glycemic control. Of note, 51.0% (51 out of 100) of non-obese LE patients on glucocorticoids, 30.6% (22/72) of obese LE patients not on glucocorticoids, and 38.6% (17/44) of obese LE patients on glucocorticoids had no HbA_1c_ value in their electronic charts. In those three groups, 17.0% (n=17/100), 47.2% (n=34/72), and 31.8% (n=14/44) of the patients had documented prediabetes or diabetes (blue and yellow in Figure 2C), suggesting that obesity is a bigger driver of dysglycemia in the LE-ASCVD Study Cohort than are GCs. For comparison, the figure for non-obese LE patients not on glucocorticoids is 23.4% (n=36/154; leftmost column in Figure 2C). For patients whose most recent HbA_1c_ value was indicative of prediabetes or diabetes, meaning ≥5.7%,[36] the time elapsed since that abnormal value was 48 months (IQR 24-80), i.e., four years (Supplementary Table S1).

For LE patients with a diagnosis of prediabetes, almost half (33/67, 49.3%) did not meet either of the two key criteria for well-managed prediabetes – namely, documentation of lifestyle modification counseling or an active prescription for metformin prescription in their charts[36–40] (Figure 2D). For LE patients with diabetes, however, over three-quarters were well-managed, generally defined as the most recent HbA_1c_ ≤7%[36–40] (n=30/39, 76.9%), and only 10.3% (4/39) had a most recent HbA_1c_ >7% (Figure 2D).

Regarding triglyceride management, only 10.0% (10/100) of non-obese LE patients on glucocorticoids, 5.6% (4/72) of obese LE patients not on glucocorticoids, and 9.1% (4/44) of obese LE patients on glucocorticoids had no plasma triglyceride value in their electronic charts (Figure 2E). Of all patients in the LE-ASCVD Study Cohort with available triglyceride values (n=331 out of 370, 89.5%), only 0.6% (2/331) had very high levels (≥500 mg/dl) and only 6.0% (20/331) had high levels (200-499 mg/dl; Figure 2E). These data indicate that hypertriglyceridemia was not a common problem in our LE-ASCVD Study Cohort even amongst patients with obesity and/or glucocorticoid therapy.

### Comparisons of three different systems for estimating the risk of future ASCVD events in LE patients

We examined three systems for estimating the risk of future ASCVD events in LE patients – namely, the newly proposed risk categories of Keyes et al, which include both primary and secondary prevention,[1] and the two widely used online risk calculators, cited above, that estimate 10-year ASCVD event risk and are limited to primary prevention. To compare these systems in our LE-ASCVD Study Cohort, we focused on the subset of patients for whom both the QRisk3 and the ACC ASCVD Risk Estimator Plus could be used, meaning sufficient data in the electronic chart and no clinically evident ASCVD (n=254 out of 370 when including age-scaling, i.e., the same number classified by the ACC ASCVD Risk Estimator Plus in Figure 1C). We separated these 254 patients into “High Risk” (n=41 out of 254; blue dots in Figure 3A and Supplementary Figure S2A) and “Very High Risk” (n=213/254; yellow dots in Figure 3A and Supplementary Figure S2B) following the categories of Keyes et al.[1] None of these 254 LE patients met criteria for the categories of “Extreme Risk” or “Recent Recurrent ASCVD Events” from Keyes et al., because both of those categories require clinically evident ASCVD, hence secondary prevention and ineligible for the QRisk3 or ACC ASCVD risk estimators.

**Figure 3.**
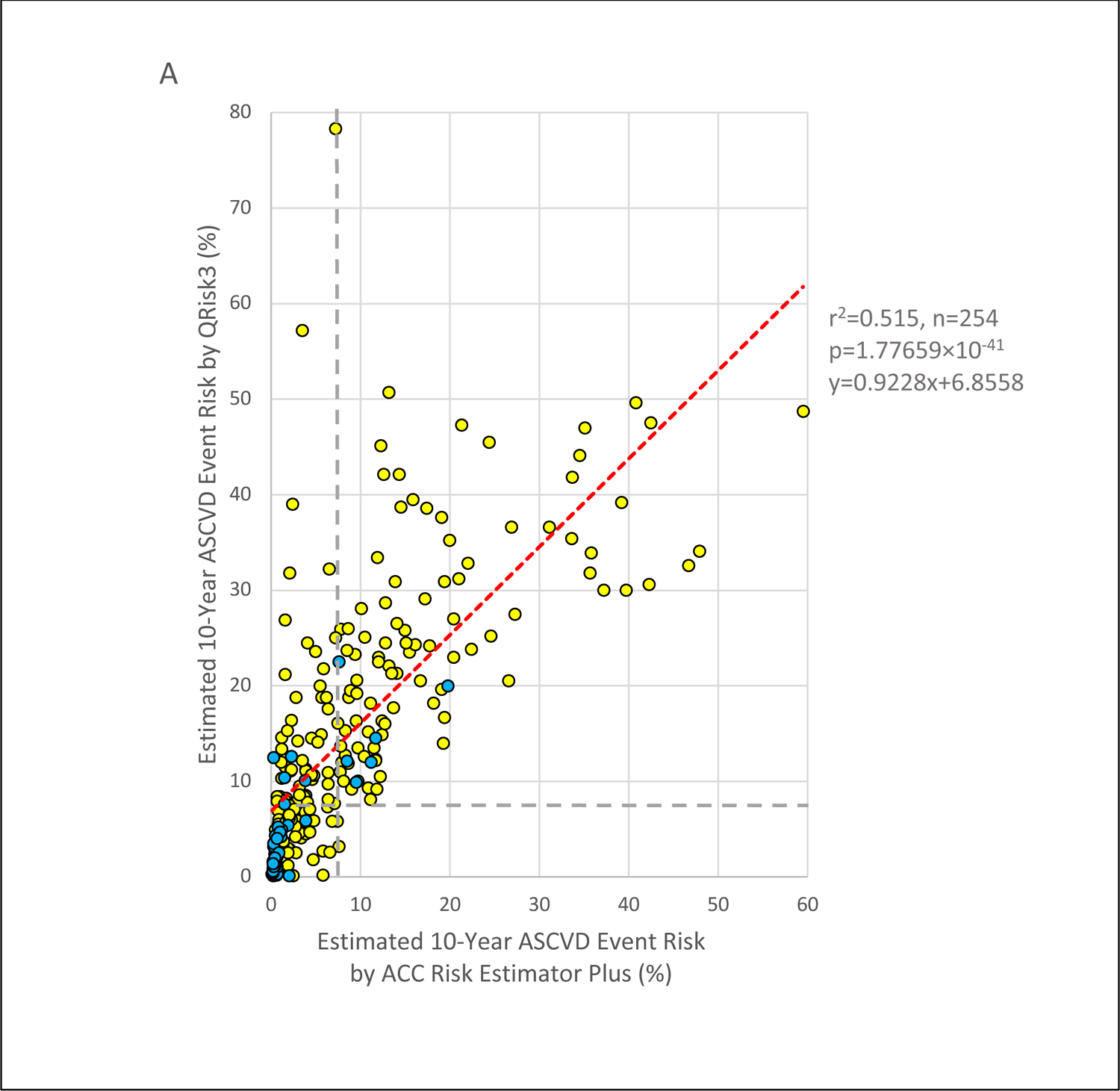
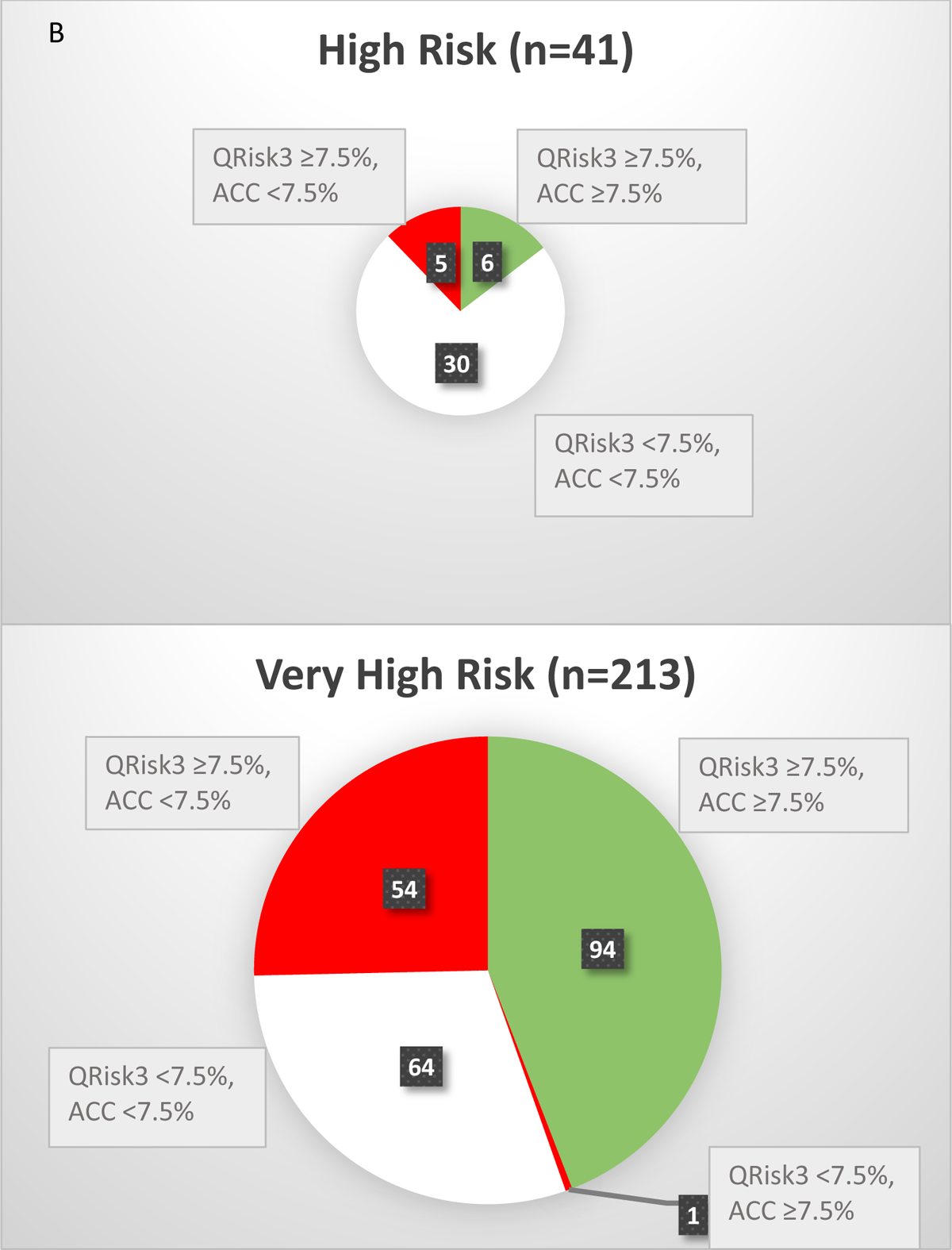
Three systems to estimate the risk of a future ASCVD event for individual patients within the LE-ASCVD Study Cohort. The three system were the ASCVD event risk categories defined by Keyes et al. for LE patients (reference[1] and Fig 1A); and numerical estimates of 10-year ASCVD event risk generated by two widely used online calculators: QRisk3 (y-axis) versus the ACC ASCVD Risk Estimator Plus (x-axis). Data are shown for the subset of patients in the Study Cohort for whom both online calculators could be used, meaning no clinically evident ASCVD (primary prevention) and sufficient data in the electronic charts to allow use of these online tools. Data are presented with (here and in Supplementary Figure S2A,B; n=254) and without (Supplementary Figure S2C,D; n=194) age-scaling for the ASCVD Risk Estimator Plus. <u>Panel A</u>: Three estimates of ASCVD event risk for each of the 254 eligible LE patients. Each dot represents an individual patient. Blue dots indicate patients classified at “High Risk” (n=41/254), and yellow dots indicate patients classified at “Very High Risk” (n=213/254) according to Keyes et al. For the two calculators, the horizontal and vertical dashed gray lines indicate the key cut-off of 7.5% estimated 10-year ASCVD event risk from the ACC/AHA guidelines for primary prevention of cardiovascular disease in the general population.[25] Pearson’s linear correlation was used to test the statistical relationship between the two numerical estimates of 10-year ASCVD event risk, and the calculated regression line is indicated in dashed red with key parameters given on the right. <u>Panel B</u>: Clinically meaningful agreement (green) and discordance (red, white) amongst the three systems for estimating the risk of a future ASCVD event for individual patients in the LE-ASCVD Study Cohort. The two pie charts tabulate LE patients at “High Risk” as defined by Keyes et al. (n=41/254) and at “Very High Risk” (n=213/254). Green indicates clinically meaningful agreement, defined here as classification of an LE patient into the “High Risk” or “Very High Risk” categories of Keyes et al., with estimates of 10-year ASCVD event risk ≥7.5% from both of the online calculators. Red indicates clinically significant discordance between the two online calculators, meaning one estimate of 10-year ASCVD event risk <7.5% but the other ≥7.5% for the same LE patient. White indicates discordance between Keyes et al. versus both of the online risk estimators, i.e., these patients are at High or Very High Risk according to Keyes et al., yet both online calculators gave estimates of 10-year ASCVD event risk <7.5%. Numbers of patients represented by each portion of each pie chart are given in the small filled black rectangles.

Figure 3A shows estimates of each of these patient’s 10-year ASCVD event risk from the QRisk3 and from the ACC calculator. Supplementary Figure S2A shows both estimates of 10-year ASCVD event risk for only the LE patients who were stratified into the “High Risk” category of Keyes et al., with the axes expanded for ease of viewing the data. Supplementary Figure S2B shows both 10-year ASCVD event risk estimates for only the LE patients stratified into the “Very High Risk” category of Keyes et al. Supplementary Figure S2B allows an unobstructed view of the data for these Very High Risk patients, indicating that many of them had remarkably low estimates of 10-year ASCVD event risk from one or both of the online calculators, a point we return to in Figure 3B.

To directly compare the numerical estimates of 10-year ASCVD event risk from the QRisk3 and the ACC ASCVD Risk Estimator Plus in our LE-ASCVD Study Cohort, we performed Pearson’s correlation with age-scaling (Figure 3A, n=254 patients) and without age-scaling (Supplementary Figure S2C, n=194 patients, i.e., the same number classified by the ACC ASCVD Risk Estimator Plus in Supplementary Figure S1C). Correlations were highly statistically significant, with p-values of 1.78×10^-41^ and 1.40×10^-27^, respectively. Nevertheless, the r^2^ values were surprisingly low, at 0.52 and 0.46, respectively. In other words, in our Study Cohort, only about half of the variation in one widely used online 10-year ASCVD risk estimate can be statistically attributed to variation in the other. Moreover, the y-intercepts were 6.9% and 7.7%, which are substantial compared with the cutoffs in the ACC/AHA guidelines for primary prevention of cardiovascular disease in the general population[25] and indicate generally higher 10-year ASCVD risk estimates from the QRisk3 than from the ACC ASCVD Risk Estimator Plus, particularly at the latter’s lower end. Visual inspection of Figure 3A and Supplementary Figures S2A-C demonstrates this point as well.

Figure 3B indicates clinically meaningful agreement (green) and discordance (red, white) amongst the three systems for estimating the risk of a future ASCVD event for individual patients in the LE-ASCVD Study Cohort. We compared the two online calculators with the newly proposed categories of ASCVD event risk for LE patients from Keyes et al. by using the key cutoff of 7.5% estimated 10-year ASCVD event risk from the ACC/AHA guidelines for primary prevention of cardiovascular disease in the general population, because estimates ≥7.5% come with substantially more aggressive recommendations for management compared with the recommendations for patients with estimates <7.5%.[25] In Figure 3B, green indicates clinically meaningful agreement, defined here as classification of an LE patient into the “High Risk” or “Very High Risk” categories of Keyes et al., with estimates of 10-year ASCVD event risk ≥7.5% from both of the online calculators. Red indicates clinically significant discordance between the two online calculators, meaning one estimate of 10-year ASCVD event risk <7.5% but the other ≥7.5% for the same LE patient. White indicates discordance between Keyes et al. versus both of the online risk estimators, i.e., these patients are at High or Very High Risk according to Keyes et al., yet both online calculators gave estimates of 10-year ASCVD event risk <7.5%. By these definitions of clinically meaningful agreement and discordance, the three systems for estimating the risk of a future ASCVD event were in agreement for only 100 of the 254 LE patients (39.4%), i.e., discordance for almost two-thirds of the LE patients (154/254, 60.6%) that could affect their clinical management. Supplementary Figure S2D shows similar findings for the 194 LE patients without age-scaling.

### Clinically documented ASCVD Events and actual 10-year ASCVD event rate in the LE-ASCVD Study Cohort

Because of discordances amongst the three systems for estimating ASCVD event *risk* (Figure 3, Supplemental Figure S2), we sought to determine the actual 10-year ASCVD event *rate* in our LE-ASCVD Study Cohort. Figure 4 shows the Kaplan-Meier curve of event-free survival, with 95% confidence intervals, since enrollment. For completeness, Supplementary Figure S3 shows the cumulative incidence of all documented MAACE in our LE-ASCVD Study Cohort, including the period before enrollment (indicated at time 0) and then all first MAACE after enrollment (time >0). Table 2 gives a breakdown of the types of MAACE.

**Figure 4.**
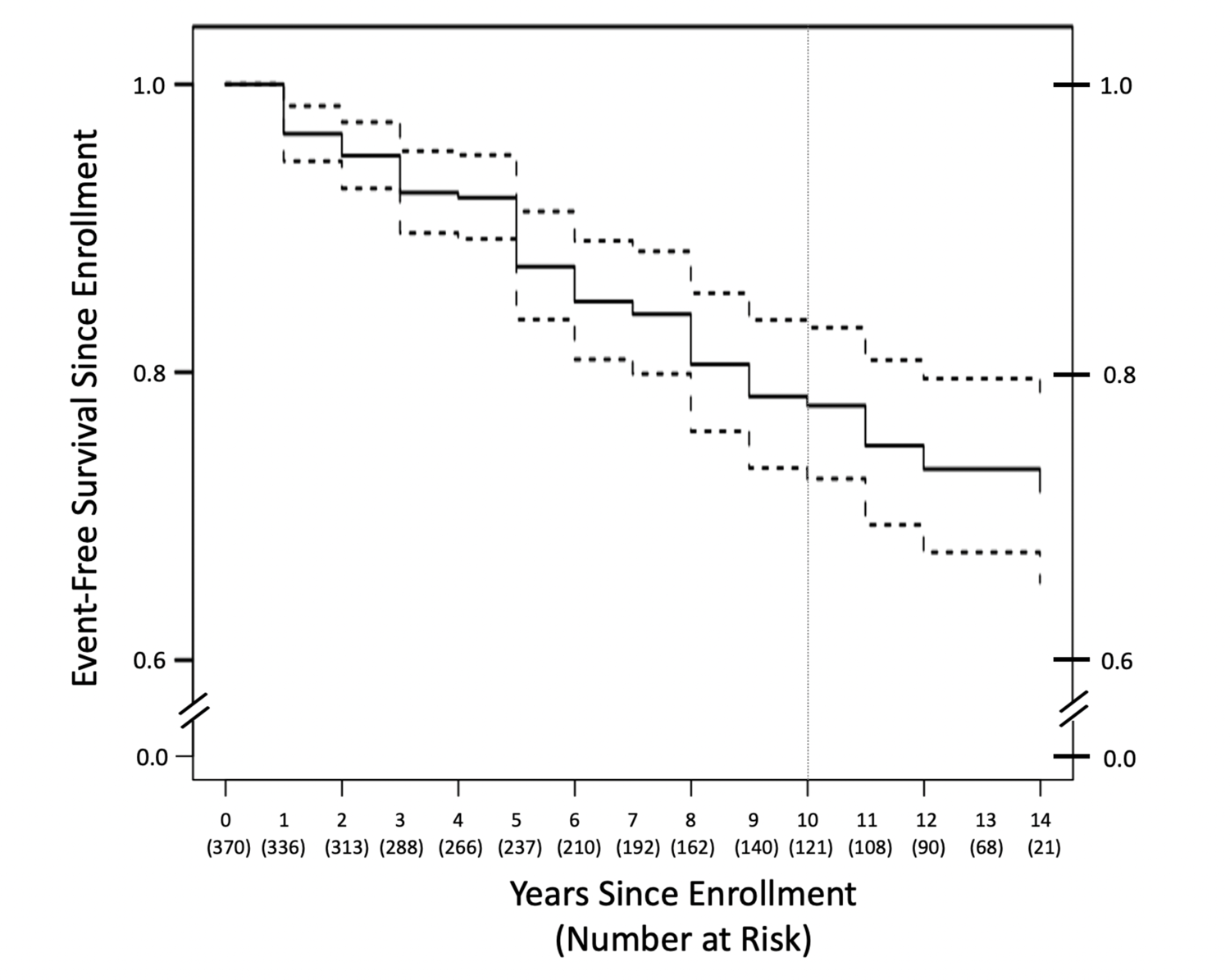
Kaplan-Meier curve of event-free survival since enrollment in the LE-ASCVD Study Cohort. An event was defined as a patient’s first documented major adverse atherosclerotic cardiovascular event (MAACE) after enrollment. Displayed are median event-free survival rates over time (solid black stepped curve) with 95% confidence intervals (dashed curves). Following our definition, the 17 MAACE that were documented before enrollment are not shown on this graph. All patients in the LE-ASCVD Study Cohort were included in this survival curve. The actual documented 10-year ASCVD event rate in our Study Cohort was calculated as one minus the event-free survival rate of 0.777 (95% CI 0.726, 0.831) at the 10-year time point, which is highlighted by the thin vertical gray line.

**Table 2.** Types, numbers, and timing of major adverse atherosclerotic cardiovascular events (MAACE) documented in the LE-ASCVD Study Cohort.

| Type of MAACE | Number affected<br>(% of the 370<br>patients in the<br>LE-ASCVD<br>Study Cohort)* | Time from enrollment<br>to event in years,<br>given as median<br>(IQR); and number of<br>events (n)‡ |
| --- | --- | --- |
| Atherosclerotic myocardial infarction (MI) | 14 (3.8%) | 6.5 (4-11); n=12 |
| Newly diagnosed ischemic heart disease other than MI | 31 (8.4%) | 8.0 (5-13); n=25 |
| Atherosclerotic (non-embolic) ischemic stroke or<br>transient cerebrovascular ischemic attack | 15 (4.1%) | 11.0 (7-14); n=10 |
| Newly diagnosed symptomatic peripheral vascular<br>disease (PVD) | 3 (0.8%) | 5.0; n=3 |
| Hospitalization for chest pain, shortness of breath, or<br>palpitations with suspicion of ischemic origin | 29 (7.8%) | 7.0 (5-8); n=28 |
| Any MAACE | 82 (22.2%) | 7.0 (5-11); n=65 |

\*Any patient who experienced two or more MAACE before and since enrollment was counted only once in any given row but was counted in two or more rows if the types of MAACE differed. Thus, the sum of the numbers of patients affected by each specific type of MAACE (92 total, 78 since enrollment) is slightly larger than the number of patients affected by any MAACE (82 total, 65 since enrollment).
‡Time and n in this column exclude the 17 events that were documented in lupus patients before their enrollment in the LE-ASCVD Study Cohort. IQR ranges are given for each type of MAACE only if n ≥ 6.

The rate of ASCVD events for patients in the first 10 years after enrollment into our LE-ASCVD Study Cohort was calculated as one minus the event-free survival at the 10-year time point of the Kaplan-Meier survival curve (Figure 4). Thus, the 10-year ASCVD event rate was 22.3% (95% CI 16.9%, 27.4%), which is above Arnett et al.’s highest cut-off of 20%,[25] indicating a high-risk population.

This 10-year ASCVD event rate in our LE-ASCVD Study Cohort is even more striking, given the preponderance of women and the patients’ relative youth at enrollment (Table 1). For comparison, the ACC ASCVD Risk Estimator Plus gives an estimate of 10-year ASCVD event risk ≥20% for a 47-year-old woman if she has a combination of diabetes, current smoking, treated hypertension, an HDLc of 40 mg/dl, and a non-HDLc ≥250 mg/dl if Caucasian and ≥160 mg/dl if African-American.

## Discussion

Our results indicate that CLE patients are under-treated compared with the new guidelines for conventional therapeutic targets to prevent ASCVD events[1] and, accordingly, these patients experience a substantial burden of major adverse ASCVD events despite our youngish, predominantly female LE-ASCVD Study Cohort. Moreover, current methods give discordant estimates of the risk for future ASCVD events, possibly because the online risk calculators were not designed specifically for CLE patients. Thus, for patients with cutaneous LE, it is unclear how to accurately assess their future ASCVD event risk – except that it is high – and this uncertainty may complicate clinical management. Management of ASCVD event risk has become a major issue in LE because of therapeutic success: lupus patients are now much less likely to die prematurely from lupus directly or from infections, and so they are living long enough to develop clinically significant atherosclerotic cardiovascular disease.[1, 6]

Specific areas of current concern include under-management of plasma LDLc, non-HDLc, and presumably apoB levels to the new guidelines;[1] under- or non-administration of LDL-lowering therapies in primary and even secondary prevention; under-management of hypertension to recently revised blood-pressure goals;[17, 34] under-use of resources for smoking cessation; infrequent monitoring of above-goal LDLc and HbA_1c_ levels; under-management of prediabetes; and, as noted above, clinically meaningful discordances in estimates of the risk for future ASCVD events and a strikingly high 10-year ASCVD event rate. Areas of current success include high levels of linkage with a PCP; evidence of widespread awareness amongst PCPs and sub-specialists of conventional therapeutic targets for management of ASCVD event risk; generally well-managed diabetes mellitus; and a low prevalence of hypertriglyceridemia in the LE-ASCVD Study Cohort even in patients with obesity and/or GC use.

Our findings add to a growing literature emphasizing the gaps between typical or usual real-world care versus ideal guideline-based care in the management of ASCVD event risk in lupus patients and in non-autoimmune populations, even ones at high risk. Several studies have looked at rates of cardiovascular events in CLE cohorts.[1, 11] To our knowledge, however, ours is the first study to focus on the management of ASCVD event risk in CLE, although there is a prior literature in SLE.[1] In one published study, a large proportion of a cohort of 110 patients with SLE had indications for statins for primary or secondary prevention, but only about half of them were treated to guidelines.[16] In a recently reported cohort of 1532 SLE patients,[17] 639 had hypertension defined by the new cutoffs of ≥130/≥80 mm Hg or on anti-hypertensive medications, but approximately three-quarters of these hypertensive SLE patients (471/639, 73.7%) were under- or un-treated, nearly identical to our figure of 74.4% (198/266, Figure 2A). Moreover, lupus patients with blood pressures in the newly defined range for stage 1 hypertension, meaning 130–139/80–89 mm Hg,[34] had more than twice the rate of atherosclerotic cardiovascular events than did normotensive lupus patients during a decade or so of follow-up.[17, 41] In other SLE cohorts as well, persistent hypertension has been associated with a much higher incidence of ASCVD events, compared with ASCVD event rates in normotensive SLE patients.[42]

In non-autoimmune populations, treatment of therapeutic targets to manage ASCVD event risk, particularly hypercholesterolemia[43–47] and hypertension,[48, 49] remains sub-optimal in the USA and worldwide. Particularly striking are under-utilization of evidence-based LDL- and blood pressure-lowering medications in patients with diabetes mellitus,[50, 51] even when those patients are at extreme ASCVD event risk owing to the additional presence of clinically evident atherosclerosis (secondary prevention) on top of their type 2 diabetes.[46]

Reasons for this widespread pattern of undertreatment might stem from prioritization of immediate clinical problems over long-term care, difficulties for patients and providers in sustaining long-term treatment for subclinical conditions, time needed to disseminate and implement recent guidelines, access to care, the difficulties of polypharmacy, evidence-based and non-evidence-based concerns from patients and providers over potential side-effects,[47, 52] and lifestyle and medication adherence. Further research is needed so that rational strategies can be developed to address under-management of ASCVD event risk. Amongst conventional therapeutic targets in the management of ASCVD event risk, levels of LDLc, non-HDLc, and apoB can be managed without great concern of a lower limit using current agents,[53, 54] in contrast to blood pressure or plasma glucose levels. Conventional interventions to assist smoking cessation and to manage prediabetes also come with few risks.

Notably, our results emphasize the need to clarify how to appropriately assess the risk of future ASCVD events in CLE patients. We demonstrated discordance amongst the three systems that we used: the new risk classifications from Keyes et al. specifically for lupus patients,[1] the ACC ASCVD Risk Estimator Plus,[24, 25] and the QRisk3 calculator.[26–28] A similar discordance between the Framingham Risk Score and QRisk3 was recently reported for SLE patients in one study[55] but less so in another study with a far smaller cohort.[16] As a statistical matter, multiplying the 10-year estimate from the ACC Risk Estimator Plus by 1.5, as has been proposed,[1, 56] would not improve the r^2^ value of the correlation with QRisk3. Recent reports suggest that QRisk3 may be a better predictor for lupus patients than other estimators of future ASCVD event risk.[57, 58] Additional clinical parameters that may enhance assessment of ASCVD event risk in lupus patients may include plasma apoB measurements,[1, 59] one-time assessments of plasma lipoprotein(a) levels, which are still largely genetically determined[1], and arterial imaging to detect subclinical plaque by iliofemoral or carotid ultrasonography or magnetic resonance imaging; coronary, iliofemoral, or carotid coronary CT angiography; and in older patients coronary artery calcium scores.[1]

### Study Limitations and Strengths

Study limitations include the retrospective design, from a single medical center. In our database, like many others, there are possible inaccuracies in smoking status because assays of cotinine, a recognized marker of nicotine use, are not part of routine care. Lifestyle modifications or medications discussed at a clinic visit but not entered into the charts could indicate a different level of management. In addition, the standard of care for diabetes testing includes assays besides HbA_1c_, such as fasting glucose levels, oral glucose tolerance tests (OGTT), and random plasma glucose levels, which may also be used to diagnose and assess prediabetes and diabetes.[36] As noted in Figure 2C, many patients in our LE-ASCVD Study Cohort had no available HbA_1c_ values, which affects our statistics on prevalence of prediabetes and diabetes and their management

Study strengths include a well-characterized longitudinal Cohort followed for over 14 years, with key parameters available in the electronic charts. The study assessed all available conventional therapeutic targets in managing ASCVD event risk, which has not been previously done in a comprehensive manner in CLE patients. Moreover, our results identified specific, key issues in clinical management and decision-making.

## Conclusions

Cutaneous LE patients are under-treated compared with the new guidelines and, accordingly, they experience a substantial burden of ASCVD events. Moreover, it is unclear how to accurately assess future ASCVD event risk in these patients – except that it is high – and this uncertainty may complicate clinical management. Based on our findings, efforts are now underway to improve ASCVD event risk estimation and guideline implementation in lupus patients.

## List of Abbreviations

ACC: American College of Cardiology

AHA: American Heart Association

apoB: apolipoprotein-B

ASCVD: atherosclerotic cardiovascular disease

CLE: cutaneous lupus erythematosus

HbA_1c_: glycated hemoglobin

HDLc: high-density lipoprotein cholesterol

Lp(a): lipoprotein(a)

LDLc: low-density lipoprotein cholesterol

LE: lupus erythematosus

MAACE: major adverse atherosclerotic cardiovascular event

MACE: major adverse cardiovascular event

MI: myocardial infarction,

OGTT: oral glucose tolerance test

PAD: peripheral arterial disease

PCP: primary care physician

SLE: systemic lupus erythematosus

TC: total cholesterol

TG: triglycerides

UPHS: University of Pennsylvania Health System

## Contributions

MZ, VPW, and KJW, were involved in the conceptualizing the project, writing the manuscript, and editing the manuscript. MZ was responsible for data collection, and MZ and KJW were responsible for data verification. MZ, RF, and KJW were responsible for data analysis.

## Data sharing

De-identified patient-level data, the full dataset, and statistical code are available from the first author (MZ). Participant consent for data sharing was not obtained but the presented data are anonymized and risk of identification is low.

## Ethics

This study was approved by the University of Pennsylvania Institutional Review Board; IRB PROTOCOL#: 849182

## Transparency Declaration

This manuscript is an honest, accurate, and transparent account of the study being reported. There are no important aspects of the study have been omitted and discrepancies have been explained. There are no competing interests.

## Data Availability

All data produced in the present study are available upon reasonable request to the authors.

