## Supplementary Materials for "THE STATE OF CURRENT MANAGEMENT OF THE HEIGHTENED RISK FOR ATHEROSCLEROTIC CARDIOVASCULAR EVENTS IN AN ESTABLISHED COHORT OF PATIENTS WITH LUPUS ERYTHEMATOSUS"

### Supplementary Materials
### for

#### THE STATE OF CURRENT MANAGEMENT OF THE HEIGHTENED RISK FOR ATHEROSCLEROTIC CARDIOVASCULAR EVENTS IN AN ESTABLISHED COHORT OF PATIENTS WITH LUPUS ERYTHEMATOSUS

Megan Zhao, BA[1,2], Rui Feng, PhD[3], Victoria P. Werth, MD[1,2*], Kevin Jon Williams, MD[4*‡]

[1]Corporal Michael J. Crescenz Veterans' Administration Medical Center, Philadelphia, PA, USA
[2]Department of Dermatology, University of Pennsylvania, Perelman School of Medicine, Philadelphia, PA
[3]Department of Biostatistics, University of Pennsylvania, Perelman School of Medicine, Philadelphia, PA
[4]Department of Cardiovascular Sciences, Department of Medicine, Lewis Katz School of Medicine at Temple University, Philadelphia, PA, USA

ORCIDs
MZ: 0000-0002-8005-9111
RF: 0000-0003-4151-7228
VPW: 0000-0003-3030-5369
KJW: 0000-0002-0000-2159

[‡]To whom correspondence should be addressed:
Kevin Jon Williams, MD
Professor of Cardiovascular Sciences, Professor of Medicine
Lewis Katz School of Medicine at Temple University
Medical Research Building (MRB)
3420 North Broad Street, Room 220
Philadelphia PA 19140
USA

**List of material included:**
Supplementary Methods
Supplementary Figures S1-S3
Supplementary Table S1

#### Supplementary Methods

*Demographic and clinical data collected for the RedCap database*

Demographic and clinical data collected for the LE-ASCVD Study Cohort included sex, race/ethnicity, age at enrollment, age as of 2021, time since enrollment in our cohort, time since CLE diagnosis, whether there was also a diagnosis of SLE, and, if so, time since SLE diagnosis. When simultaneous heights and weights were available, we calculated body mass indices (BMIs), to stratify the patients into categories of underweight (commonly defined as <18.5 kg/m$^2$ but alternatively as <20.0 kg/m$^2$ based on mortality data[60]), normal weight (commonly defined as 18.5 to 24.9 kg/m$^2$ but also as 20.0 to 24.9 kg/m$^2$ [60]), overweight (25.0 to 29.9 kg/m$^2$), or obese (≥30.0 kg/m$^2$). Amongst the LE-ASCVD Study Cohort, a diagnosis of depression in the electronic medical record, history of lupus nephritis, history of chronic kidney disease (CKD) stage 3 or higher, and smoking status (never, former, or current) from chart abstraction were also entered into our RedCap database.

In our RedCap database, we also included whether there was a listed primary care provider (PCP), and the date of the last PCP visit was noted. All specialties caring for the patients within the past 20 years, if available, were compiled.

Orders and results for lipid/lipoprotein panels and glycated hemoglobin ($HbA_{1c}$) levels were extracted from the electronic medical records, including dates and the specialty of the ordering physician.

All blood pressure values in each patient's chart were entered into our RedCap database, to allow us to assess recent blood pressure management and to calculate blood pressure variability, a parameter included in the QRisk3 calculator. To diagnose hypertension, we averaged blood pressure readings from the most recent year of follow-up.

Documentation in the electronic charts of counseling on lifestyle changes, specifically diet, exercise, weight loss, and smoking cessation, was noted along with dates of counseling. Referrals to dieticians, weight-loss programs, and smoking cessation clinics were also included. From physicians' and other providers' notes and the medication lists in the charts, we extracted prescriptions for medications for lipid-lowering, hypertension, and glycemic control, as well as the dates of these prescriptions.

*Three systems to estimate the risk of a future ASCVD event for individual patients within the LE-ASCVD Study Cohort*

Based on these clinical data, including absence (primary prevention) or presence (secondary prevention) of clinically evident ASCVD, as well as major conventional risk factors for ASCVD events, we classified patients from the LE-ASCVD Cohort into the four defined categories of ASCVD event risk from our recently proposed guidelines for ASCVD event risk management for LE patients – namely, High Risk, Very High Risk, Extreme Risk, and Recent Recurrent ASCVD Events.[1] Next, we compared current clinical care with the new therapeutic goals for each of these categories of ASCVD event risk for LE patients.[1]

In addition, we evaluated patients in the LE-ASCVD Cohort with two widely used online calculators that estimate 10-year ASCVD event risk for individuals from the general population, i.e., not specifically designed for CLE patients. Use of these online calculators was limited to patients with sufficient data in the electronic chart and no clinically evident ASCVD (primary prevention); the calculators are not designed to estimate future ASCVD event risk in patients who have already had an atherosclerotic cardiovascular event (secondary prevention).

One of the online calculators that we used was the ACC ASCVD Risk Estimator Plus, which may be the best known but does not take a diagnosis of LE into consideration.[24, 25] Moreover, it calculates 10-year ASCVD event risk only for patients aged 40-79 years. To allow calculations as broadly as possible, patients in our LE-ASCVD Cohort below age 40 years without clinically evident ASCVD for whom we had sufficient data for the ACC ASCVD Risk Estimator Plus were scaled up to 40, and patients above age 79 years without clinically evident ASCVD were scaled down to 79 (referred to as "age-scaling" in the main Results section and legends to figures and supplementary figures). For simplicity, we did not include multipliers of this estimation of 10-year ASCVD event risk, although some authors have advocated such multipliers for LE patients.[1, 56] There were just two patients with TC >320 mg/dL, the highest value allowed by the ACC ASCVD Risk Estimator Plus, so we scaled those values to 320. We stratified LE patients by their estimated 10-year ASCVD event risks from the ACC Risk Estimator Plus, following the cutoffs of Arnett et al. in the ACC/AHA guidelines for primary prevention of cardiovascular disease in the general population[25] – namely, 0.0-4.9% 10-year ASCVD event risk (low), 5.0-7.4% (borderline), 7.5-19.9% (intermediate), and ≥20% (high).

The other online calculator that we used to estimate 10-year ASCVD event risk was QRisk3, which includes SLE status and use of orally administered steroids, but not CLE status, and requires that patients not be prescribed statins at baseline.[26, 27] QRisk3 can be used for patients aged 25-84 years, so we scaled up three patients to an age of 25 and scaled down four patients to an age of 84. The QRisk3, which is based in the United Kingdom, draws on data for specific racial/ethnic groups there that do not include large numbers of African Americans or Hispanics, and so patients from our LE-ASCVD Cohort in those demographics had to be classified under the QRisk3 category of "Other ethnic group". The ACC ASCVD Risk

Estimator Plus, which is based in the USA and therefore more aligned with the population of our catchment area, does not have this limitation. Online estimates were performed by one author (MZ), and divergent estimates of 10-year event risk between the two online calculators were spot-checked by another author (KJW).

#### Supplementary Figures and Table

**Supplementary Figure S1.** Assessment and management of plasma non-high-density lipoprotein cholesterol (non-HDLc) and LDLc concentrations in the LE-ASCVD Study Cohort. Panel A: Classification of Study Cohort patients into the four categories of ASCVD event risk defined by the newly proposed guidelines of Keyes et al. for lupus patients.[1] Also indicated are numbers of patients with non-HDLc levels above (red) or within (green) the newly proposed goals for each risk category. Patients without an available non-HDLc value (or no simultaneous total cholesterol and HDLc) are indicated in blue. Panel B: Absence (red) or presence (green) of an active prescription for an LDL-lowering medication in the electronic charts of patients classified into each of the four newly proposed categories of ASCVD event risk for lupus patients and whose most recent plasma non-HDLc levels were above the newly proposed goals; these are the patients indicated in red in panel S1A. Panel C: Estimated risk of an ASCVD event in the next 10 years ("10-year ASCVD event risk") by the online ACC Risk Estimator Plus and absence (red) or presence (green) of an active prescription for an LDL-lowering medication in the electronic chart. Patients are grouped according to the cut-offs for estimated 10-year ASCVD event risk from the ACC/AHA guidelines for primary prevention of cardiovascular disease in the general population (0.0-4.9%, 5.0-7.4%, &c).[25] As noted in the Supplementary Methods and in Figure 1, patients below age 40 years without clinically evident ASCVD for whom we had sufficient data for the ACC ASCVD Risk Estimator Plus (n=47/370) were scaled up to 40, and patients above age 79 years without clinically evident ASCVD (n=13/370) were scaled down to 79. Data are presented with these additional patients included (Figure 1C) or presented separately (here). Also shown here are data for patients with clinically evident ASCVD (secondary prevention, for which the ACC ASCVD Risk Estimator Plus does not apply), as well as data for patients with no clinically evident ASCVD but insufficient information in the electronic chart for the ACC Risk Estimator (rightmost column). Panel D: Most recent LDLc values and absence (red) or presence (green) of an active prescription for an LDL-lowering medication in the electronic chart. The rightmost column represents patients with no LDLc value in the electronic chart. Panel E: Most recent non-HDLc values and absence (red) or presence (green) of an active prescription for an LDL-lowering medication. The rightmost column represents patients with no non-HDLc value (or no simultaneous total cholesterol and HDLc) in the electronic chart. At the top of each column in panels A-E is indicated the total number of patients represented by that column. Numbers of patients represented by each tinted portion of each column are also given.

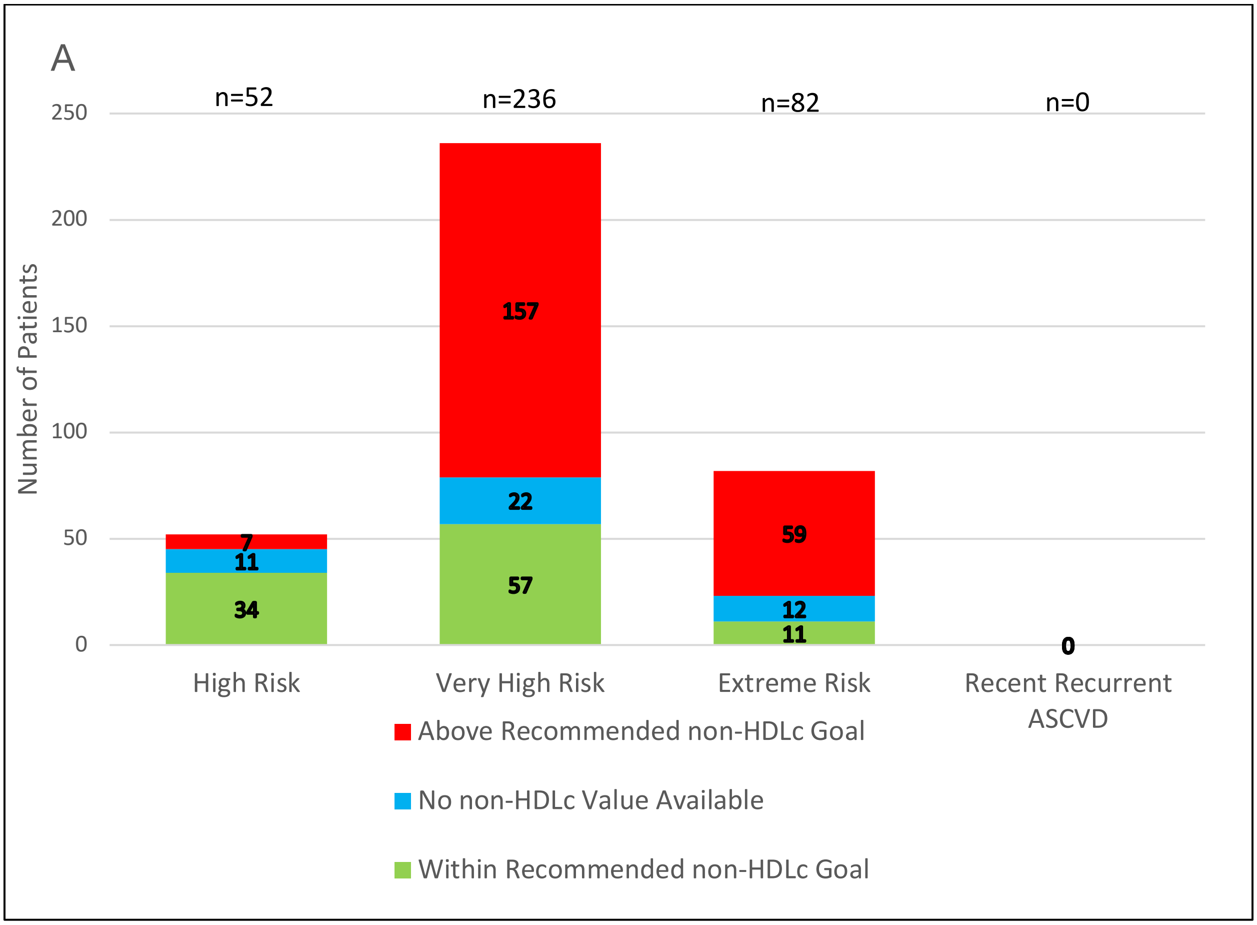
A
n=52
n=236
n=82
n=0
250
200
150
100
50
0
Number of Patients
157
22
57
7
11
34
59
12
11
0
High Risk
Very High Risk
Extreme Risk
Recent Recurrent ASCVD
Above Recommended non-HDLc Goal
No non-HDLc Value Available
Within Recommended non-HDLc Goal

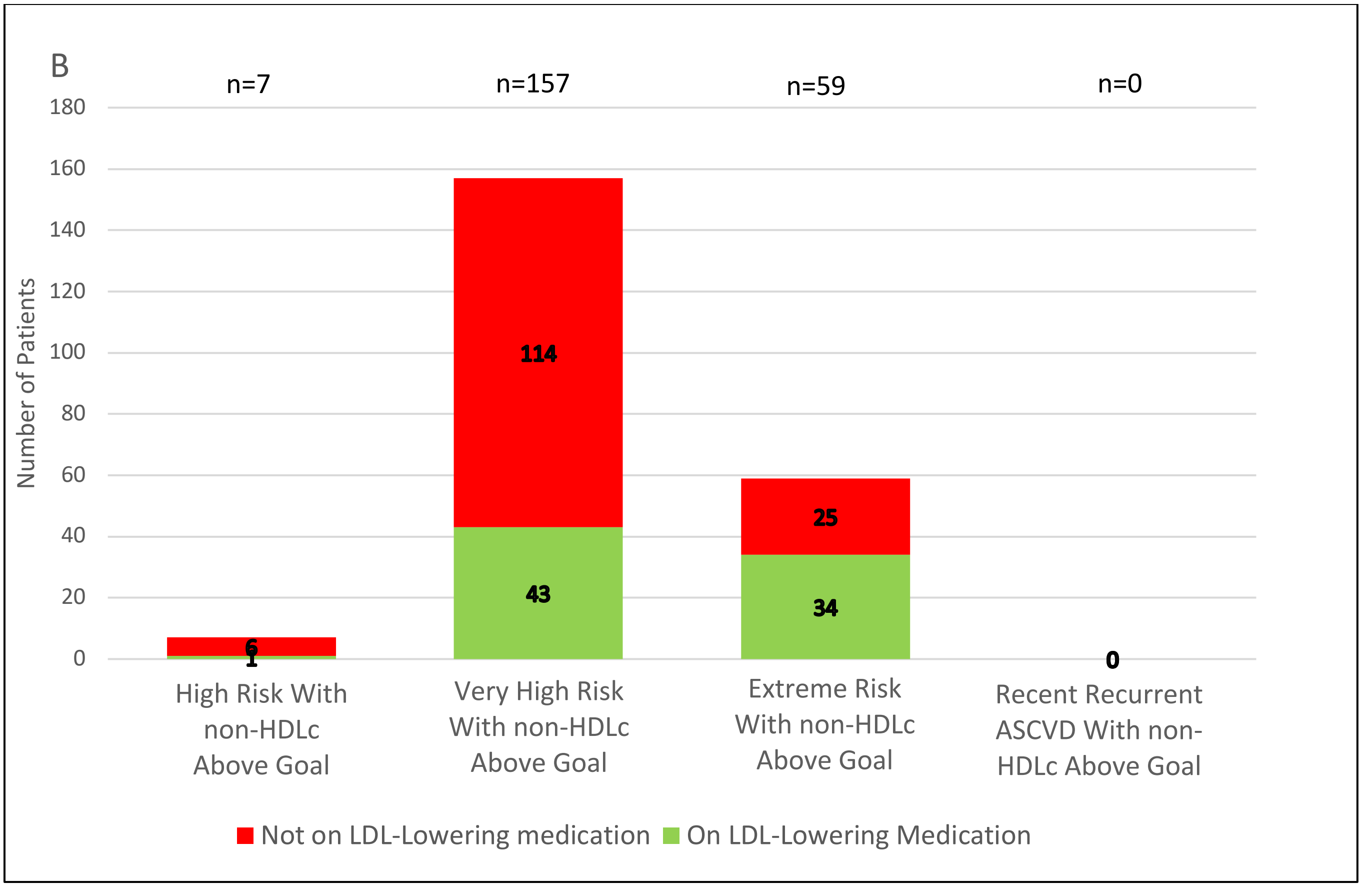
B
n=7
n=157
n=59
n=0
Number of Patients
180
160
140
120
100
80
60
40
20
0
6
1
114
43
25
34
0
High Risk With non-HDLc Above Goal
Very High Risk With non-HDLc Above Goal
Extreme Risk With non-HDLc Above Goal
Recent Recurrent ASCVD With non-HDLc Above Goal
Not on LDL-Lowering medication
On LDL-Lowering Medication

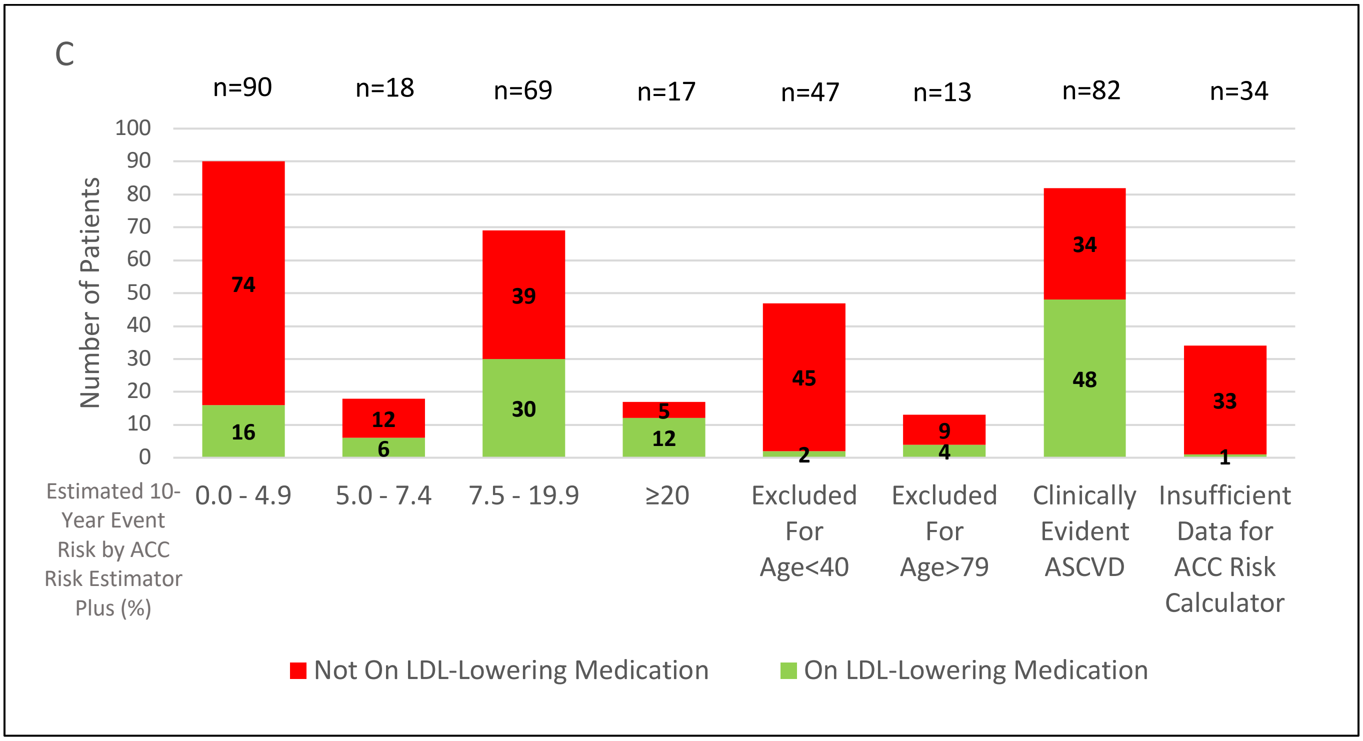
C
n=90
n=18
n=69
n=17
n=47
n=13
n=82
n=34
Number of Patients
100
90
80
70
60
50
40
30
20
10
0
74
16
12
6
39
30
5
12
45
2
9
4
34
48
33
1
Estimated 10-Year Event Risk by ACC Risk Estimator Plus (%)
0.0 - 4.9
5.0 - 7.4
7.5 - 19.9
≥20
Excluded For Age<40
Excluded For Age>79
Clinically Evident ASCVD
Insufficient Data for ACC Risk Calculator
Not On LDL-Lowering Medication
On LDL-Lowering Medication

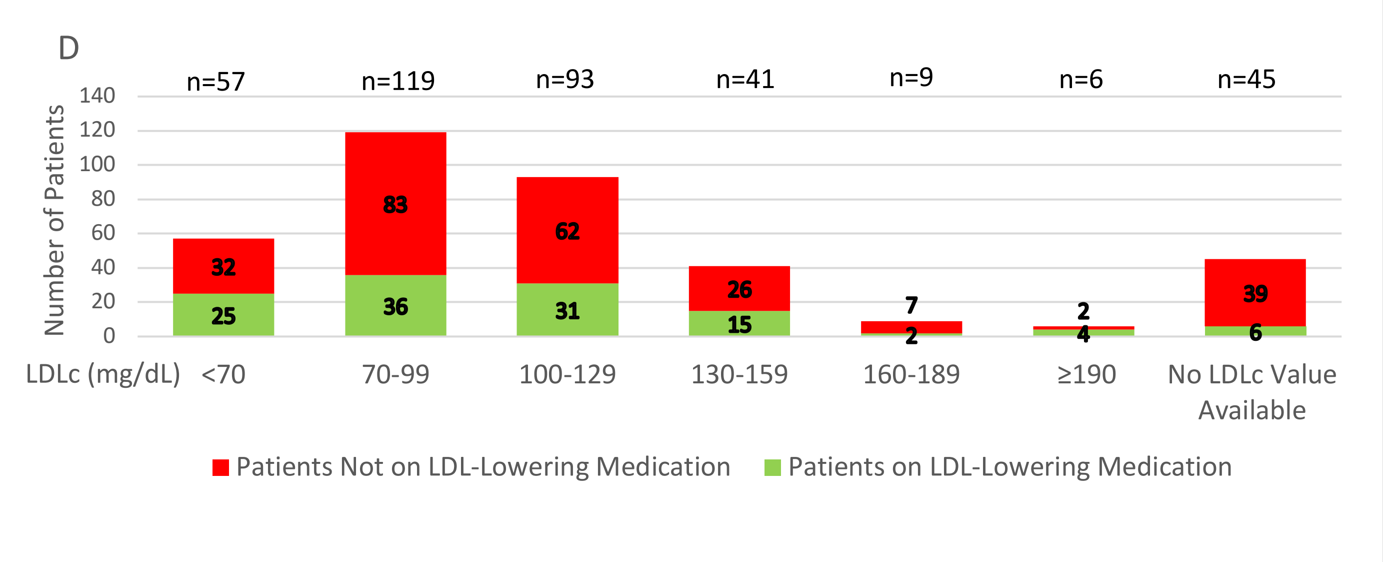
D
Number of Patients
0
20
40
60
80
100
120
140
n=57
n=119
n=93
n=41
n=9
n=6
n=45
32
25
83
36
62
31
26
15
7
2
2
4
39
6
LDLc (mg/dL)
<70
70-99
100-129
130-159
160-189
≥190
No LDLc Value Available
Patients Not on LDL-Lowering Medication
Patients on LDL-Lowering Medication

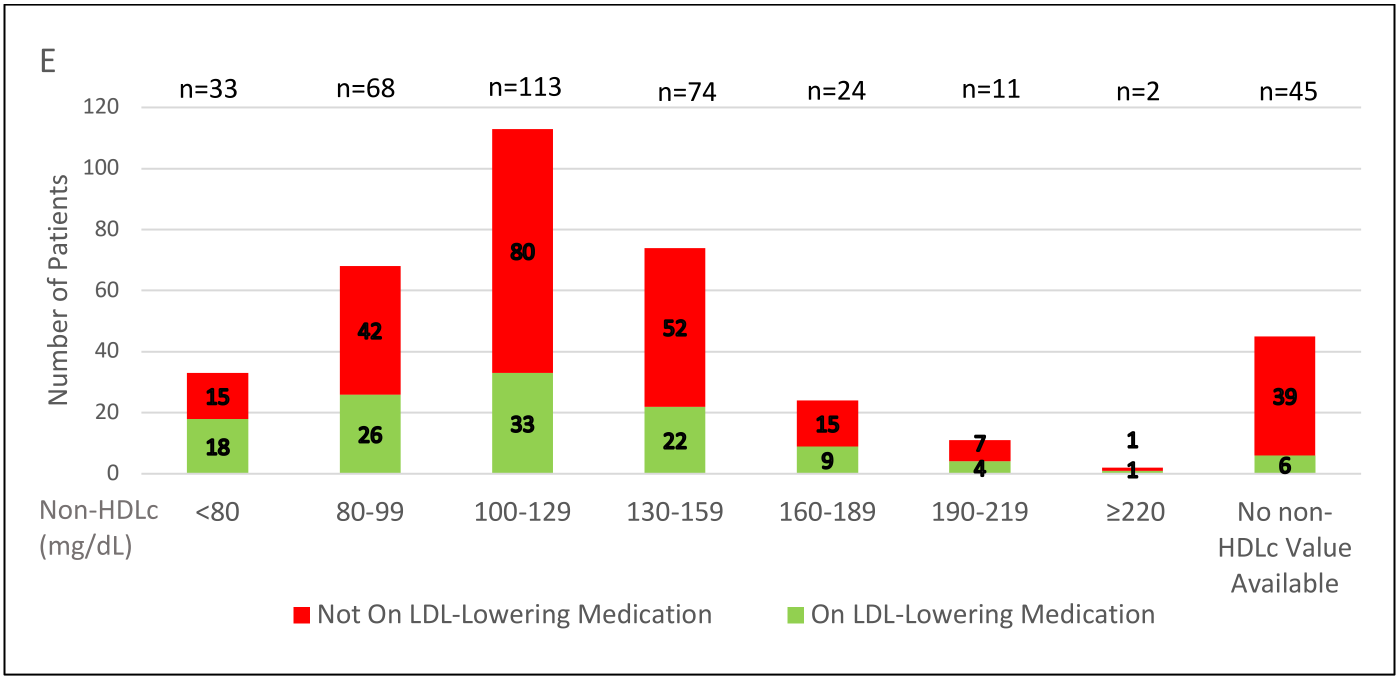
E
Number of Patients
0
20
40
60
80
100
120
n=33
n=68
n=113
n=74
n=24
n=11
n=2
n=45
15
18
42
26
80
33
52
22
15
9
7
4
1
1
39
6
Non-HDLc (mg/dL)
<80
80-99
100-129
130-159
160-189
190-219
≥220
No non-HDLc Value Available
Not On LDL-Lowering Medication
On LDL-Lowering Medication

**Supplementary Figure S2.** Three systems to estimate the risk of a future ASCVD event for individual patients in the LE-ASCVD Study Cohort, as described in Figure 3. Data are shown for the subset of patients in the Study Cohort for whom both online calculators could be used, meaning no clinically evident ASCVD (primary prevention) and sufficient data in the electronic charts to allow use of these online tools. Symbols and labeling in these panels follow Figure 3. Panel A: Estimates of 10-year ASCVD event risk, with age-scaling, for LE patients classified into the "High Risk" category (n=41/254) of Keyes et al.[1] These data are also shown in Figure 3A, but the panel here has been expanded for ease of viewing the data. Panel B: Estimates of 10-year ASCVD event risk, with age-scaling, for LE patients classified into the "Very High Risk" category (n=213/254) of Keyes et al. These data are also in Figure 3A, but the panel here allows an unobstructed view of the data for these Very High Risk patients. Panel C: Presentation of data with (Figure 3A and panels A,B here; n=254) and without (panels C,D here; n=194) age-scaling for the ACC ASCVD Risk Estimator Plus. Here, patients classified at "High Risk" were 31/194 (blue dots), and "Very High Risk" were 163/194 (yellow dots). As in Fig. 3A, Pearson's linear correlation was used to test the statistical relationship between the two numerical estimates of 10-year ASCVD event risk, and the calculated regression line is indicated in dashed red with key parameters given on the right. Panel D: Clinically meaningful agreement (green) and discordance (red, white) amongst the three systems for estimating the risk of a future ASCVD event for individual patients in the LE-ASCVD Study Cohort. This panel is same as Figure 3B except that it does not include patients whose ages were scaled to allow use of the ACC ASCVD Risk Estimator Plus.

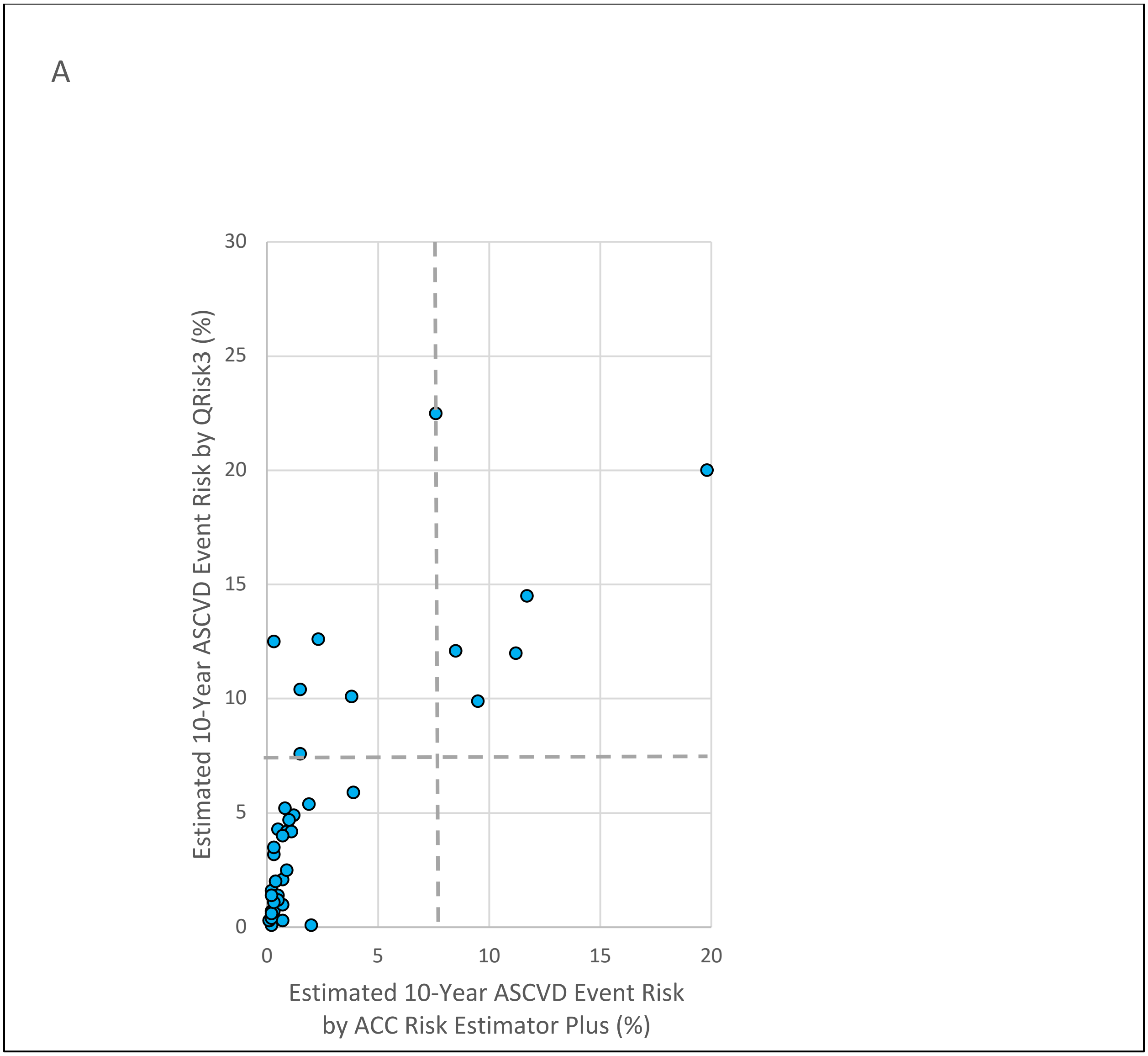
A
30
25
20
15
10
5
0
0
5
10
15
20
Estimated 10-Year ASCVD Event Risk by QRisk3 (%)
Estimated 10-Year ASCVD Event Risk
by ACC Risk Estimator Plus (%)

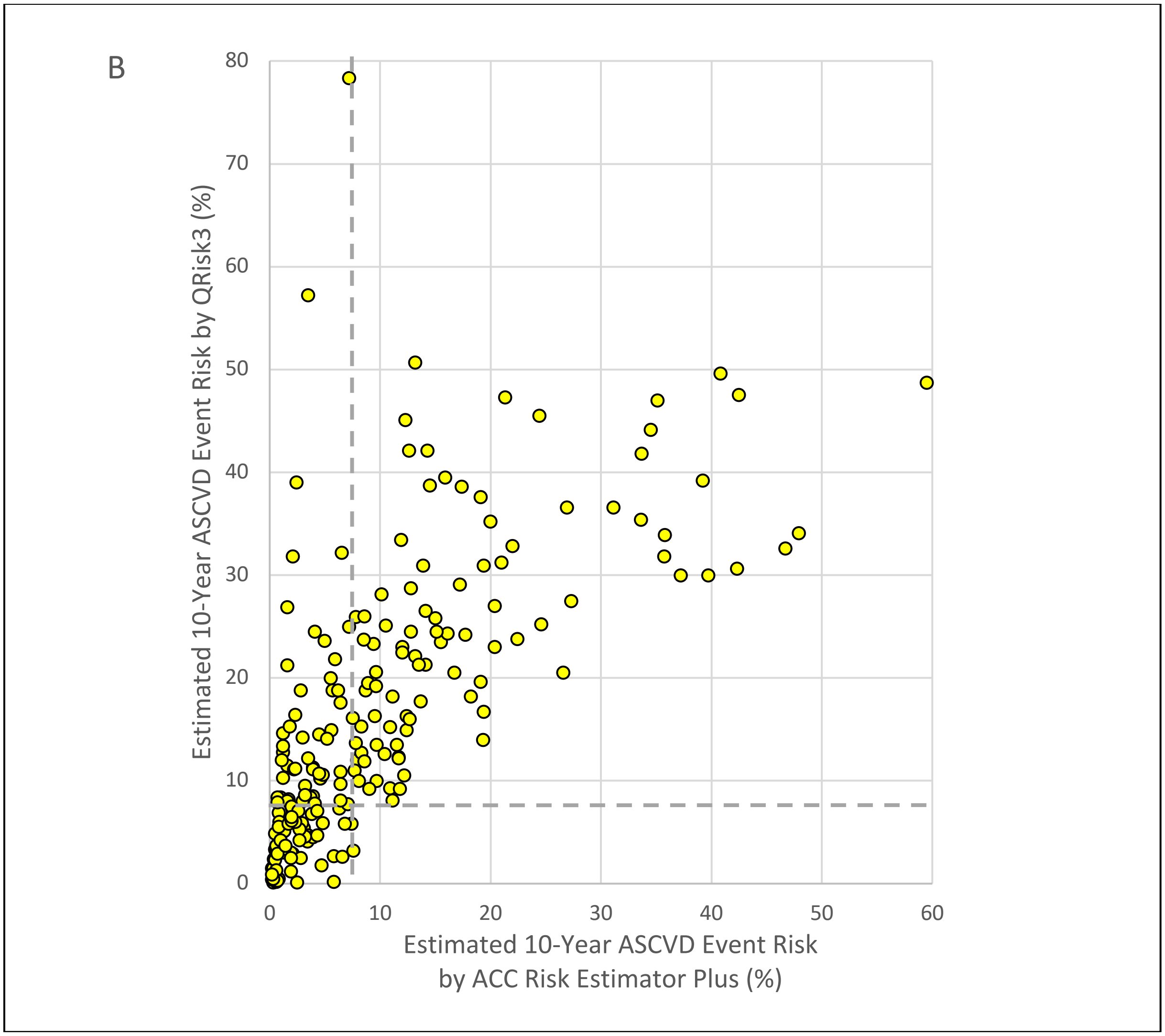
B
80
70
60
50
40
30
20
10
0
0
10
20
30
40
50
60
Estimated 10-Year ASCVD Event Risk by QRisk3 (%)
Estimated 10-Year ASCVD Event Risk
by ACC Risk Estimator Plus (%)

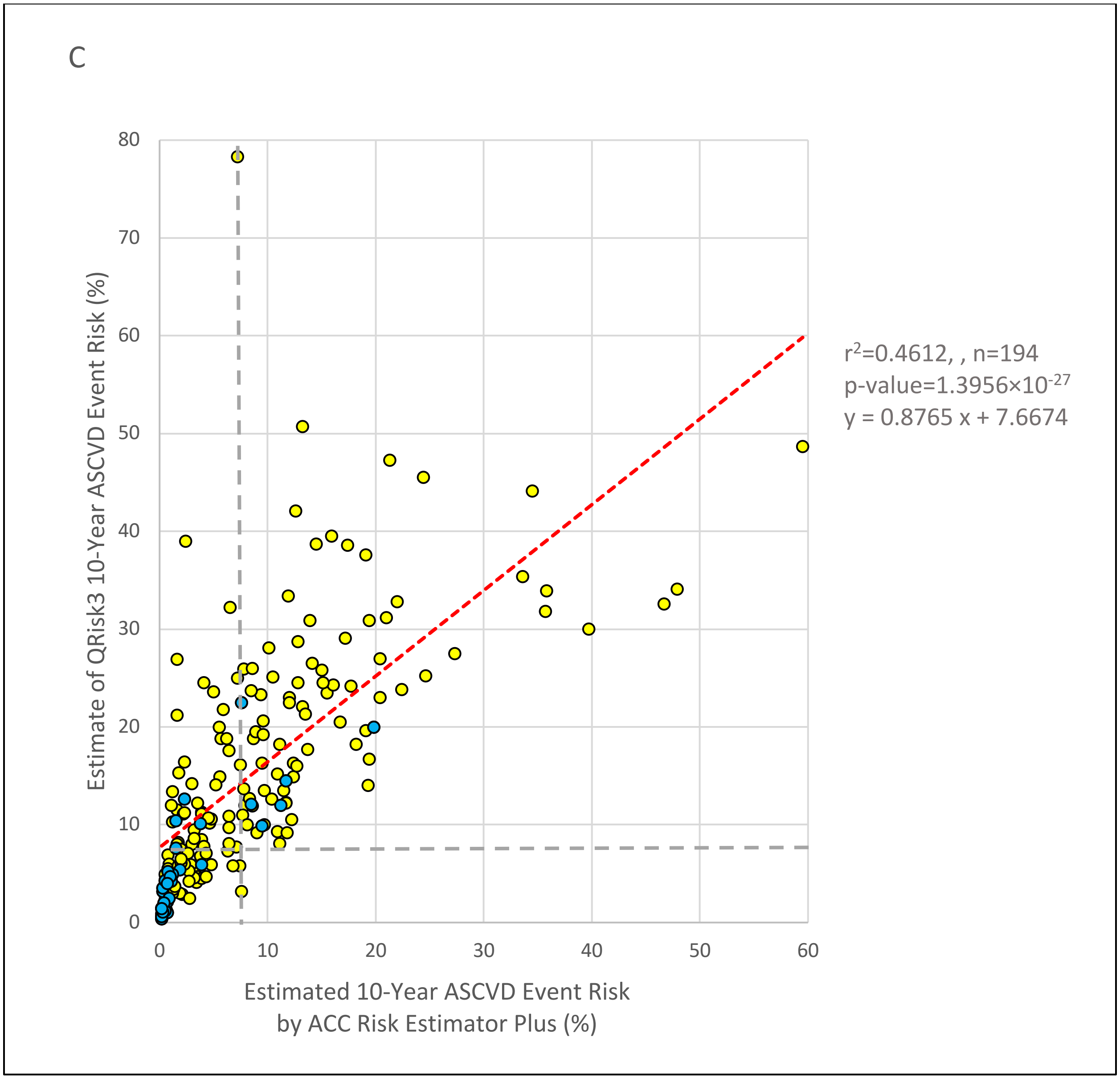
C
Estimate of QRisk3 10-Year ASCVD Event Risk (%)
80
70
60
50
40
30
20
10
0
0
10
20
30
40
50
60
Estimated 10-Year ASCVD Event Risk
by ACC Risk Estimator Plus (%)
$r^2$=0.4612, , n=194
p-value=1.3956×$10^{-27}$
y = 0.8765 x + 7.6674

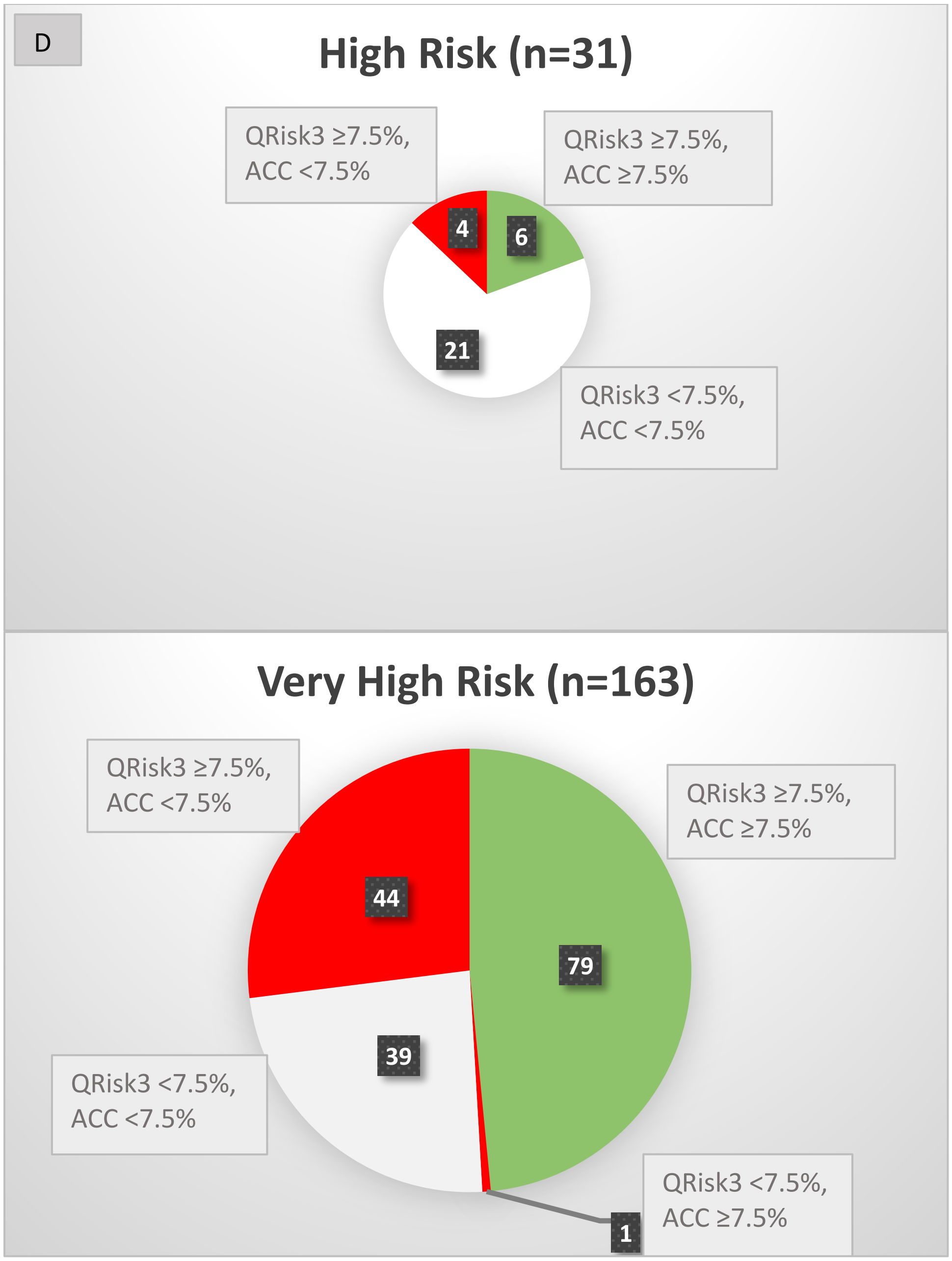
D
High Risk (n=31)
QRisk3 ≥7.5%, ACC <7.5%
QRisk3 ≥7.5%, ACC ≥7.5%
QRisk3 <7.5%, ACC <7.5%
4
6
21
Very High Risk (n=163)
QRisk3 ≥7.5%, ACC <7.5%
QRisk3 ≥7.5%, ACC ≥7.5%
QRisk3 <7.5%, ACC <7.5%
QRisk3 <7.5%, ACC ≥7.5%
44
79
39
1

**Supplementary Figure S3.** Cumulative incidence of documented major adverse atherosclerotic cardiovascular events (MAACE) in the LE-ASCVD Study Cohort. The 17 events that were documented before enrollment in the LE-ASCVD Study Cohort occurred in 17 lupus patients and are indicated at time 0 (n=17/370, 4.59%). For all patients, the first MAACE after enrollment was counted for this curve (time >0) but no subsequent MAACE. After enrollment, the numbers of patients in the Cohort year by year are consistently higher than the numbers at risk in the Kaplan-Meier curve of event-free survival (Figure 4), because when a patient has a non-fatal event, he or she becomes censored from the K-M curve, but still remains in the Cohort.

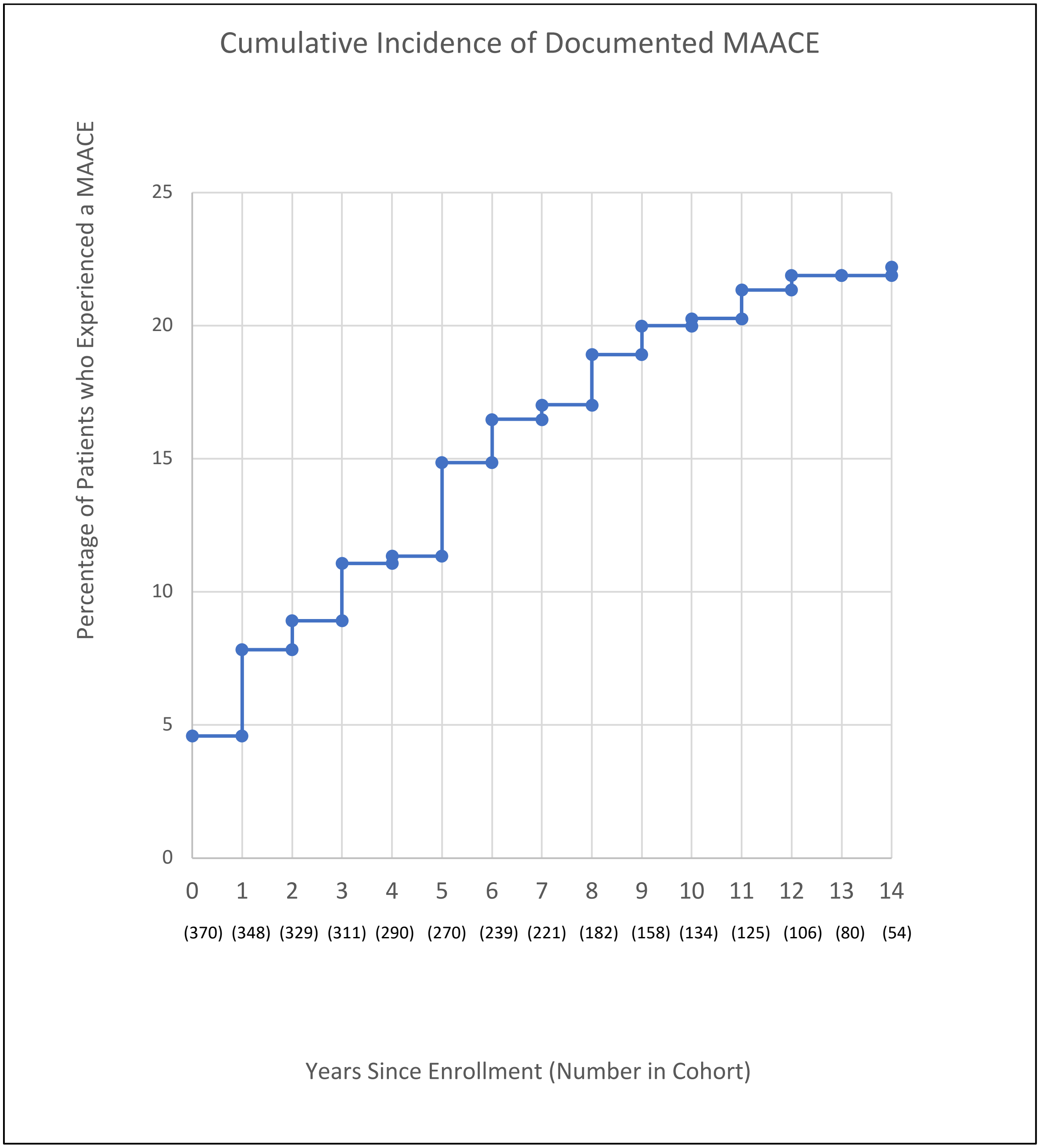

**Supplementary Table S1.** Additional characteristics of the LE-ASCVD Study Cohort. Categorical parameters are given as n (%). Continuous variables are given as mean±SD, if normally distributed, or as median (interquartile range, IQR), if non-normally distributed.

| **Category** | **Parameter** | **Value** | **n in LE-ASCVD Study Cohort or subgroup** |
|---|---|---|---|
| **Additional clinical characteristics** | | | |
| | Time since diagnosis of cutaneous LE | 13.0 years (IQR 8.25-21.0) | 370 (entire LE-ASCVD Study Cohort) |
| | Time since diagnosis of cutaneous LE | 15.0 years (IQR 10.0-25.0) | 190 (subgroup of CLE patients who have concurrent SLE) |
| | Time since diagnosis of systemic LE | 13.0 years (IQR 8.00-23.8) | 190 (same subgroup as above) |
| | Underweight (BMI <18.5 kg/m$^2$, common definition) | 13 (3.8%) | 346 (subgroup with at least one set of simultaneous height and weight measurements) |
| | Underweight (BMI <20.0 kg/m$^2$ based on mortality data[60]) | 21 (6.1%%) | 346 |
| | Normal weight (BMI 18.5 to 24.9 kg/m$^2$, common definition) | 112 (32.4%) | 346 |
| | Normal weight (BMI 20.0-24.9 kg/m$^2$ based on mortality data[60]) | 104 (30.1%) | 346 |
| | Overweight (BMI 25.0 to 29.9 kg/m$^2$) | 105 (30.3%) | 346 |
| | Diagnosis of depression in the electronic chart | 139 (37.6%) | 370 |
| **Additional data regarding providers** | | | |
| | Time since last visit with PCP | 10 months (IQR 4.0-31) | 279 (subgroup with at least one PCP visit documented in the chart) |

| | | | |
|---|---|---|---|
| | Number of physician providers, excluding trainees | 5.8±2.8 (mean±SD) | 370 |
| | Patients who have a recorded visit with a rheumatologist | 203 (54.9%) | 370 |
| | Patients who have a recorded visit with a cardiologist | 151 (40.8%) | 370 |
| | Patients who have a recorded visit with a nephrologist | 57 (15.4%) | 370 |
| | Patients who have a recorded visit with an endocrinologist | 33 (8.0%) | 370 |
| **Assessments of plasma lipids/lipoproteins** | | | |
| | Lipids/lipoproteins checked at least once | 325 (87.8%) | 370 |
| | Time since the most recently documented lipid/lipoprotein assay | 35 months (IQR 14-83) | 325 (subgroup with at least one lipid/lipoprotein result in the chart) |
| | PCP checked the most recent lipids/lipoproteins | 204 (62.8%) | 325 |
| | Non-PCPs checked the most recent lipids/lipoproteins | 121 (37.2%) | 325 |
| | • Rheumatology | 35 (28.9%) | 121 (subgroup for whom non-PCPs checked the most recent lipids/lipoproteins) |
| | • Cardiology | 15 (12.4%) | 121 |
| | • Nephrology | 15 (12.4%) | 121 |
| | • Endocrinology | 3 (2.5%) | 121 |
| | • Emergency | 19 (15.7%) | 121 |
| | • Dermatology | 15 (12.4%) | 121 |
| | • Other | 19 (15.7%) | 121 |
| | Time elapsed since above-goal LDLc value | 47 months (IQR 24-97) | 252 (subgroup of patients whose most recently documented LDLc |

| | | | |
|---|---|---|---|
| | | | value in the chart was above the newly recommended goals, i.e., red in Figure 1A) |
| **Assessments of levels of glycated hemoglobin ($HbA_{1c}$)** | | | |
| | $HbA_{1c}$ checked at least once | 208 (56.2%) | 370 (entire LE-ASCVD Study Cohort) |
| | Time elapsed since the most recently documented $HbA_{1c}$ assay | 36 months (IQR 11-68) | 208 (subgroup with at least one $HbA_{1c}$ value in the chart) |
| | PCP checked the most recent $HbA_{1c}$ | 132 (65.3%) | 202 |
| | Non-PCPs checked the most recent $HbA_{1c}$ | 70 (34.7%) | 202 |
| | • Rheumatology | 18 (25.7%) | 70 (subgroup for whom non-PCPs checked the most recent $HbA_{1c}$) |
| | • Cardiology | 14 (20.0%) | 70 |
| | • Nephrology | 2 (2.9%) | 70 |
| | • Endocrinology | 3 (4.3%) | 70 |
| | • Emergency | 12 (17.1%) | 70 |
| | • Dermatology | 9 (12.9%) | 70 |
| | • Other | 12 (17.1%) | 70 |
| | Time elapsed since abnormal $HbA_{1c}$ value | 48 months (IQR 24-80) | 89 (subgroup of patients whose most recently documented $HbA_{1c}$ value in the chart was abnormal, meaning ≥5.7%) |
